# Field-ready portable rapid nucleic acid test for tuberculosis detection and drug-resistance profiling in resource-limited settings

**DOI:** 10.64898/2026.05.29.26354438

**Authors:** Sudip Nag, Saptarshi Banerjee, Sohom Banerjee, Subham Ghosh, Arijit Bera, Sivakumar Shanmugam, Arindam Mondal, Suman Chakraborty

## Abstract

Tuberculosis (TB) remains one of the world’s deadliest infectious diseases, with over a million deaths annually and a growing threat from multidrug-resistant strains (MDR-TB). A major bottleneck in controlling TB is the lack of truly portable, rapid, and user-friendly diagnostic systems that can operate effectively in decentralized, resource-constrained settings. Here, we present a first-of-its-kind, portable nucleic-acid-based diagnostic platform that enables both primary TB screening and detection of drug resistance within the same unified framework, without any change in the operative embodiment. The system integrates loop-mediated isothermal amplification (LAMP) targeting dual Mycobacterium tuberculosis markers (IS6110 and IS1081) with a compact, AI-enabled device and smartphone-based readout, delivering rapid and reliable results at the point-of-care. Clinical evaluation across 105 samples demonstrated high sensitivity and specificity. Further validation through real-world deployment in a primary healthcare setting, using a single-gene (IS6110) configuration operated by minimally trained personnel, yielded 95.60% sensitivity and 100% specificity, benchmarked against GeneXpert. Critically, the same platform architecture, without modification, extends seamlessly to drug-resistance profiling, demonstrated here through a probe-free, allele-specific LAMP approach for identifying key mutations associated with rifampicin (rpoB) and isoniazid (katG) resistance. By combining robust molecular diagnostics with AI-driven automation in a compact and accessible format, this work represents a significant medical advancement toward democratizing TB care. The platform thus holds strong potential to enable early screening, guide timely treatment decisions, reduce transmission, and substantially strengthen global TB elimination efforts, particularly in high-burden, low-resource settings.

## INTRODUCTION

Tuberculosis (TB) continues to rank among the deadliest infectious diseases, claiming 1.23 million lives in 2024 alone [1]. Caused by *Mycobacterium tuberculosis* (MTB), it disproportionately affects low- and middle-income countries, where limited access to diagnostics fuels ongoing transmission. The challenge is further compounded by the rise of multidrug-resistant TB (MDR-TB) or extensively drug-resistant TB (XDR), driven by genetic mutations that render standard antibiotics such as rifampicin (RIF) or/and isoniazid (INH) ineffective [2, 3]. In 2024, worldwide 1,47,592 people were detected with MDR/RR-TB and 25,140 people with pre-XDR-TB or XDR-TB, giving a combined total of 1,72,732 drug resistant TB cases, thus posing a formidable barrier to global eradication efforts [1,3].

Although the World Health Organization has set the ambitious goal of ending TB by 2035 [1], progress has been slowed by a fundamental bottleneck: timely and accessible diagnosis. Traditional culture-based tests combined with drug susceptibility testing (DST), while definitive, take up to two months and require advanced laboratories and skilled technicians’ conditions rarely available in high-burden regions [4,5]. Molecular tests like GeneXpert and Truenat [6,7] have improved detection, but their reliance on costly cartridges, trained operators, and uninterrupted electricity limits their reach in rural and resource-limited areas, often restraining their long-term sustainability in the low- and middle-income countries with a high TB burden [8,9]. Line probe assays offer resistance profiling but are too complex and slow for routine frontline use [6,10].

Loop-mediated isothermal amplification (LAMP) has emerged as a promising alternative, capable of amplifying DNA at a constant temperature through simple readouts without dependency on the thermal cycling equipment [11–13]. This technology relies on a strand displacement chemistry driven by Bst DNA polymerase enzyme and employs 4-6 primers targeting 6-8 region of the DNA template to generate distinctive cauliflower-like structures, facilitating rapid exponential amplification of the target DNA. The amplification reaction could be monitored through simple pH indicator based colorimetric readout and dye or metal ion indicator based fluorometric readout, which could be readily recoded by simple spectrophotometer/fluorimeter for quantitative results [12–14]. In principle, this technology could provide rapid, low-cost, and field-friendly TB testing. Yet despite encouraging laboratory results, its impact in real-world settings has remained elusive, hampered by design challenges, risks of contamination, lack of multiplexing, and the absence of integrated, user-friendly devices tailored for point-of-care deployment [15–19]. Only in recent times, a few attempts have been initiated in developing LAMP based rapid point-of-care testing (POCT) technologies for tuberculosis diagnosis [20–23].

In this study, we present a next generation TB diagnostic that bridges these gaps of POCT technologies for tuberculosis detection. We developed a dual-marker based LAMP assay targeting conserved genomic sequences (IS6110 and IS1081), combined with a probe-free allele-specific assay to identify resistance-conferring mutations towards rifampicin and isoniazid. Importantly, these innovations were embedded into an AI-enabled, portable, pre-programmable device which has been designed for the frontline use, bringing sophisticated molecular testing into the hands of community health workers. By integrating scientific innovation with practical usability, this platform represents a major step toward accessible, field-deployable diagnostic for TB and MDR-TB. The outstanding sensitivity (95%) and specificity (100%) of the proposed system in clinical and field evaluation strengthen this claim. It offers a realistic path to scale community screening, reduce transmission, and accelerate global efforts to end the TB epidemic. The overview of portable device, smartphone app and overall testing procedure has been depicted in Fig.1.

**Fig. 1.**
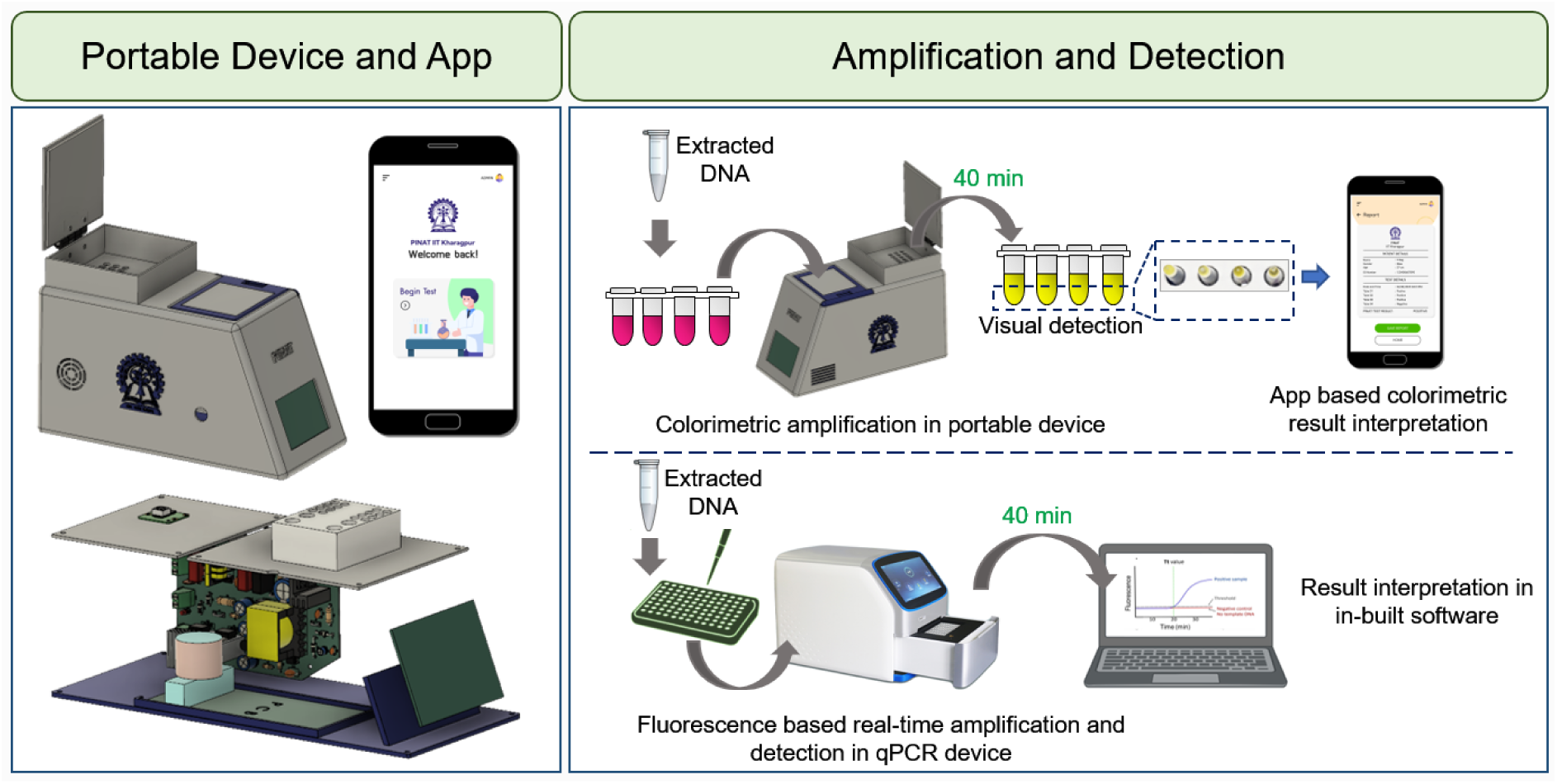
Overview of procedural steps including amplification and detection for colorimetric and real-time diagnosis of TB infections or disease. Colorimetric detection was performed by portable device and smartphone application. Fluorescence based detection was performed in real-time machine.

## MATERIALS AND METHOD

### Development of portable device, smartphone application and integration of method

An upgraded version of previously reported [13] diagnostic platform was designed and developed using Autodesk Fusion, integrating both thermal and optical modules into a compact, portable enclosure. It was included a Raspberry Pi Zero 2 W microcontroller that manages dual aluminium heating cartridges, a DS18B20 one-wire temperature sensor, and a fan-heat sink assembly, all controlled via an optocoupler relay and a switch-mode power supply. The design allowed this device to automatically execute time-stamped isothermal colorimetric protocol in a sequential manner including amplification and detection operations. The reaction unit of the device was programmed to operate at 65°C for up to 120 min and 95°C for 5 min so that the amplification process of colorimetric LAMP assay could be performed seamlessly. The thermal block has the necessary number of slots to hold an equivalent number of reaction tubes so that multiple test reactions could be operated simultaneously and the block maintained precise temperature control through a PID (Proportional-Integral-Derivative) algorithm, similar to that used in other handheld thermal devices, ensuring stability within ±0.9 °C. A visual indicator provided real-time status updates. Additionally, the system was housed in a top-loading, light-tight black enclosure that prevents ambient light interference and contains a 5 MP camera with synchronized white LED illumination for standardisation of illumination and camera quality. Reaction tubes were inserted from the top, the hinged lid was closed to create light isolation. Then the device could connect seamlessly to a smartphone interface using secure Wi-Fi, allowing users to remotely initiate image capture via an API call from the smartphone app which was developed using React Native, a cross-platform framework for Android and iOS. The app provides user functions such as registration, login, viewing test histories, uploading image and displaying result.

### App-based image capture and ML classification

The images once captured by the inbuilt camera within the device, gets automatically transferred to the smartphone application, retrieved and organised with timestamped filenames and sent it to a backend server. The algorithm pre-processed images using OpenCV, focusing on each tube’s region of interest (ROI) and converting it to HSV colour space to ensure consistent hue analysis. A support vector machine (SVM), implemented with scikit-learn, classifies each reaction as positive, negative, or uninterpretable based on average hue, saturation, and value. The ML classifier was initially trained using a labelled dataset of reaction images (approx. 1000) obtained from controlled experiments including confirmed positive reactions (yellow), negative controls (pink), and borderline reactions. These data set was divided into training and validation subsets to optimize the decision boundary and evaluate classification performance. To address potential ambiguity in color interpretation, reactions exhibiting intermediate signal intensity were classified as “uninterpretable” when feature values fell within the predefined uncertainty region between positive and negative decision thresholds. Such results were flagged by the application and recommended for repeat testing. The application was modular and optimised for inference with TensorFlow Lite, supporting batch processing, result logging, and integration with external databases. By embedding training data-driven machine learning-based interpretation in the server, the system removed subjective visual assessments, providing quick, reliable diagnostics with minimal user training, meeting the REASSURED criteria for effective point-of-care molecular testing.

### Development of the assay system for screening TB infection

Herein, simple pH based colorimetric LAMP reaction has been developed and optimized to screen TB infection and disease. The reaction system included with 1X WarmStart Colorimetric LAMP Master Mix (New England Biolabs, USA), 1X primer mix (containing six primers) and plasmid template. The colorimetric isothermal process was optimized through a specific thermal cycle as heating at 65°C from 0 min to 60 min (amplification step) and at 95°C for 5 min (termination step) which was performed in the mentioned portable device. Once the thermal protocol was completed images of the tubes were captured by device-integrated camera, sent it to the smartphone and subsequently analyzed by the app as mentioned earlier. Finally, product formation was verified by 1% agarose gel electrophoresis.

Besides, the real-time fluorometric LAMP reaction system (fluorescent metal indicator dye based) was developed to screen TB infection/disease and optimized on QuantStudio™ 5 Real-Time PCR Detection System (Thermo Fisher Scientific, USA) through the same thermal cycle as used in colorimetric LAMP. This assay system included with WarmStart non-colorimetric LAMP 2X Master Mix, 1X primer mix, of 1X Calcein dye (New England Biolabs, USA) and plasmid templates. Fluorescence of Calcein was measured in every 30 sec up to 60 min. Amplification was analyzed based on the threshold time (Tt) values normalized to the negative control (NC) threshold time. In each case, at least three independent replicates of each reaction were carried out. In each experiment, was used as the negative control (NC) in place of the positive samples.

### Development of assay system for detecting drug resistant TB infection

We also optimized an assay system for detecting the presence of drug resistant TB based on allele specific colorimetric and fluorometric LAMP reaction. The allele specific colorimetric and real-time fluorometric reaction were performed in a similar way as optimized for detecting TB infection/disease with minor necessary modification viz. the time of incubation at 65°C was extended up to 100 min. Once the colorimetric process was completed, the analysis and confirmation of outcome were carried out in a manner akin to that of the TB-LAMP assay. The real-time amplification was monitored on QuantStudio™ 5 Real-Time PCR Detection System by measuring the fluorescence in every 30 sec up to 100 min. The allele-specific LAMP assays were developed and evaluated in this study primarily as a proof-of-concept to demonstrate analytical discrimination of common resistance-associated mutations of rifampicin and isoniazid. For discriminating wild type (WT) and mutant type (MT), the reaction was carried out with two primers specific to the WT and MT alleles. Depending on screening outcomes, allele specific primers were used having mismatches either at position-1 only or both at −1, and −2, of forward inner primer’s (FIP) 3’ end position. A known concentration of WT and MT plasmids (mutations at rpoB D435V, rpoB H445D, rpoB S450L and katG S315T) positive controls of the rpoB and katG genes were used for validating the assay. Detection process of rpoB D435V, rpoB H445D, rpoB S450L and KatG S315T with respective WT was performed simultaneously in separate reaction tubes. As per assay concept, amplification implies the presence of MT allele only whereas no amplification infers the presence of WT allele, which will ultimately discriminate the drug resistant TB infections form the drug-susceptible TB infections.

### Analytical sensitivity, assay precision, specificity and receiver operating characteristic

The gene wise (IS6110 and IS1081) analytical sensitivity of colorimetric and fluorometric screening test was evaluated using different concentration (5 to 1×10^6^ copies/µL) of plasmid from respective inserts. A foresaid testing protocols were performed followed by analyzing the color difference between positive and negative samples at the end of experiments. Each experiment was conducted three times. Gene wise assay precision was assessed by testing three replicates in the same run in different days and by different operators at the 10 to 1×10^6^ copies/µL range to calculate assay variability by means of the coefficient of variance (CV). To determine the molecular specificity of colorimetric and fluorometric screening test for active form TB infection or disease, was evaluated using 15 ng of genomic DNA template extracted from other bacteria, including *Escherichia coli* (ATCC 25922), *Staphylococcus aureus* (ATCC 25923) and *Bacillus subtilis* (ATCC 6633) which was three times higher than MTB genomic DNA. Receiver Operating Characteristic (ROC) was further investigated to estimate the efficacy, based on the sensitivity and specificity of the *in-vitro* test.

### Clinical samples, validation and ethics statement

The initial validation study was performed at ICMR-NIRT, Chennai, India, which serves as a WHO collaborating centre for TB research and training and also acts as a supranational reference laboratory for mycobacteriology. A total of identified 105 genomic DNA (including both infection positive and negative), extracted from the cultures inoculated with sputum isolates, were used in this study to evaluate the overall clinical performance of the assay at laboratory set-up. Genomic DNA was extracted using automated nucleic acid extraction procedure by designated laboratory personnel of ICMR-NIRT and finally stored in their institutional repository. These samples were then subjected to TB-LAMP assay and CBNAAT (GeneXpert) assay parallelly in a double blinded manner. CBNAAT (GeneXpert) was used as the reference comparator or molecular gold standard test method.

Additionally, field performance of the test was evaluated against a cohort of 119 samples collected during a span of 60 days. Sputum samples were obtained from the suspected symptomatic individuals as well as from some healthy persons visiting at Shivshankar Pote Hospital, Pune Municipal Corporation, India. During the collection of sputum samples from each individual, we followed the guidelines and inclusion/exclusion criteria as revealed by the ICMR, India and WHO. The testing kits were prepared at IIT Kharagpur and transported to the testing centre which is around 1700 kilometres away. Special attention was provided on several factors, e.g. packaging of reaction mixtures in the form of test kits to supply at the testing centre, storage of the pre-packed kits, reagent stability under different temperature conditions, and execution of the tests exclusively at the resource-limited settings by minimally trained personnel. The details of their training have been provided in supporting information. All samples were collected from patients/volunteers with their informed consent as per the approval of the respective institutional ethical committee. The study was also approved by the IIT Kharagpur institutional ethical committee. The TB-LAMP assay and CBNAAT (GeneXpert) were tested in parallel. Both assays were performed on aliquots derived from the same clinical specimen. For TB LAMP assay, genomic DNA was extracted from the sputum samples using HiPurA® Mycobacterium tuberculosis DNA Purification Kit (Himedia, India).

Plasmid DNA containing respective target sequence (IS6110 or IS1081) at a concentration of 1000 copies/µL were used as positive control for each batch of 96 test reaction kit. RanseP gene was used as “internal control” which was included for each clinical sample. A single negative control was (nuclease free water) was included for each test run. To minimize bias, the personnel performing the assay were blinded to the CBNAAT(GeneXpert) results at the time of testing. The outcomes of screening test were eventually compared with the CBNAAT (GeneXpert) data of the test samples to determine the diagnostic performance of the assay.

### Bioinformatics analysis

For designing primers of targeted genes for screening assay and drug resistance assay, multiple sequence alignment was performed with the more than 1000 sequences using the MAFFT server (https://mafft.cbrc.jp/alignment/server/). Then, the Bio-Edit multiple sequence editor (www.mbio.ncsu.edu/bioedit/bioedit.html) was used to analyze the conservation. Conserved target regions corresponding to the particular gene were selected from the consensus sequence obtained from the multiple sequence alignment. CIRCOS maps were generated by Python using Matplotlib (https://matplotlib.org/).

### Statistical analysis

Clinical data were analysed in Microsoft Excel (V 2021). Gene wise sensitivity (positive percent agreement), specificity (negative percent agreement) and overall accuracy were calculated as per the following:

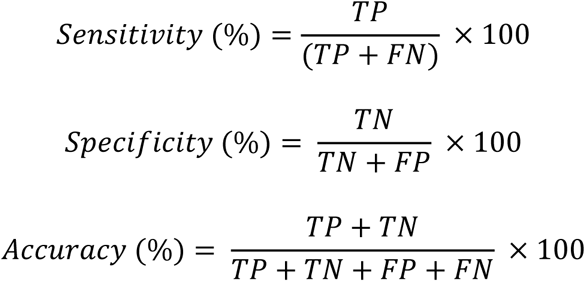

where, TP: True positive, TN: True negative, FP: False positive, FN: False negative 95% confidence interval (CI) was calculated for the data set as per previous reports [24–26]. In addition, Cohen’s Kappa (κ) has also been calculated to measures the agreement between two diagnostic methods (Index test vs Reference test) as per previous reports [27,28]. The formula has been provided in supporting information.

## RESULTS AND DISCUSSION

### IS6110 and IS1081 as dual biomarkers for screening TB infection

The Mycobacterium tuberculosis complex (MTBC), comprised of six strains are generally classified into high and low insertion elements copy number strains [29], although the effect of this genetic variation upon bacterial pathogenesis remains unclear. It was reported that the M. tuberculosis Beijing/W lineage, with an unusually high copy number of IS6110, is associated with higher virulence and massive spread of drug resistant strains [30]. Upon in-silico analysis of fully sequenced and assembled MTBC genomes, we observed that the *M. bovis* AF2122 strain contains a single copy of IS6110 whereas M. tuberculosis HN306 and *M. tuberculosis* H37Rv have high copy numbers (20 and 16) of this element (Fig. S1A). Besides, we also observed that except *M. canettii* CIPT140010059 strain, other strains harbour an average 6 copies of IS1081 elements (Fig.S1B). The insertion elements content in MTBC genomes suggested that the copy number of this transposon might be lineage-specific. Among the human-adapted species of MTB, these high copy number strains show high prevalence across the world, hence reflecting their capability of infecting and causing disease across different demographics. We analysed the possible genetic variations within these insertion elements to finalise the target regions (Table S1 and Table S2) and subsequently, design LAMP primers to amplify regions in the insertion elements, IS6110 and IS1081, that are completely conserved across all MTBC strains. While the IS6110 amplified region remained completely conserved (Fig.S1C), the IS1081 amplified region contains a single mutation (Fig. S1D) exclusively in *M. bovis* AF2122 strains (1 mutation at 490 nucleotide position). However, no genetic polymorphism was present at any of the primer annealing sites for both of the IS elements. The extensive bioinformatics analysis in determining the target regions paved the way forward towards the development of a highly robust amplification method ensuring high sensitivity and specificity of the screening test.

Notably, the intended clinical application of this assay is routine programmatic diagnosis of human pulmonary TB, for which *M. tuberculosis sensu stricto* represents the overwhelming majority of cases. Herein, the two markers were evaluated independently at the analytical and clinical levels to assess their complementary roles in detecting MTB infection. In addition to the IS6110, which has been reported as a marker in previous studies, our method included the IS1081 as an additional conserved marker, to improve the assay inclusivity and robustness in the context of IS6110 low-copy number MTBC strains as analysed (e.g. M. bovis stain contains one copy IS6110 and six copies IS1081), and to provide extra precaution against possible target failure.

### In-vitro standardization of the TB-LAMP assay

Following *in-silico* analysis, distinct set of primers for each of the IS elements were employed for *in-vitro* assay standardization. In principle, the reaction process starts with outer primers (F3, B3) initiating strand displacement, followed by inner primers (FIP, BIP) that extend and displace strands to form dumbbell shaped structures. Loop primers (LF, LB) accelerate cycling by annealing to single-stranded loops, producing enormous DNA via autocatalytic reactions without thermal cycling [14,31]. We performed the colorimetric-LAMP in the inhouse developed portable device and real-time fluorometric LAMP using the commercial RT-PCR machine in order to have a comparative analysis between them. In colorimetric test, LAMP positive amplification lead to the color change from pink to yellow due to the presence of pH-sensitive dye in mastermix. Here, positive amplification lead to the production of protons and subsequent drop in pH that occurs from the extensive DNA polymerase activity, which finally makes the reaction solution acidic. The color changes in tube was captured by the camera integrated within the portable device and subsequently interpreted using the synchronized smartphone application. The fluorescent metal indicator dye based fluorometric test was documented through real-time amplification curve using the commercial RT-PCR device. Herein, during amplification, calcein yields strong fluorescence through replacement of the resident manganese ions with the magnesium ions generated during the amplification process [14]. All reactions are also validated through the typical ladder-like pattern of the amplified products in agarose gel electrophoresis (Fig. S2).

Analytical sensitivity is the lowest DNA/RNA concentration that a test can detect without any ambiguity, thereby denotes the limit of detection (LoD) for individual target genes. For colorimetric LAMP assay, the device detected distinct color changes (pink to yellow) for the MTB genomic DNA (extracted from a clinical sample) within 40 minutes (Fig. S3). Therefore, 40 min was considered as the optimal duration for LAMP based isothermal amplification (65^0^C) of MTB DNA. Under these optimized conditions, different concentrations (hence different copy number) of individual plasmid harboring either of the IS elements were utilized to obtain the LoD for both colorimetric and fluorescence methods. The colorimetric assay consistently detected a 100 copies/µL of the gene block harboring either of the insertion elements independently (Fig. 2A). Although 10 copies/µL was also detectable for most of the trials, but the results remained inconsistent. Herein, the smartphone app successfully analyzed the received images of positive and negative outcomes and subsequently interpreted the results based on the color change. Orange coloration was also observed in limited number of amplified reactions having lower concentrations of target (beyond 100 copies/ µL), which was defined here as ‘uninterpreted’ result. The ‘uninterpretable’ results were excluded from the performance calculations. In case of the real-time LAMP assay, fluorescence sigmoidal curves were obtained at various concentrations, and the threshold time (Tt: time required to obtain the signal beyond the threshold cutoff value) showed inverse correlation with target concentrations (Fig. 2B and Fig. 2C). The threshold time (Tt) values were found to exhibit a linear correlation (R^2^ > 0.98) with the logarithmic concentrations over a wide dynamic range from 10^6^ copies/ µL to 10 copies/ µL (Fig. 2D). In both targets, the lowest detectable concentration was 10 copies/ µL. We further conducted both colorimetric and fluorometric sensitivity analysis with the genomic DNA (isolated from patient samples) and determined the LoD to be 1 ng (Fig. 2E), a value that can be easily attained with the standard DNA extraction methods used for clinical samples. It is worth mentioning that the LOD for both the colorimetric and fluorescent LAMP assays achieved here are better than the recently reported different detection methods [12, 32–34]. We also tested for specificity against other bacteria causing common hospital-acquired infections in order to assess the possibility of false positive results in actual clinical settings. As displayed in Fig. 2F (upper panel), no signals were detected from other bacterial DNA except MTB DNA within 40 min. For colorimetric test as shown in Fig.2F (lower panel), only the MTB DNA provide a detectable color change while other strains show negative results.

**Fig. 2.**
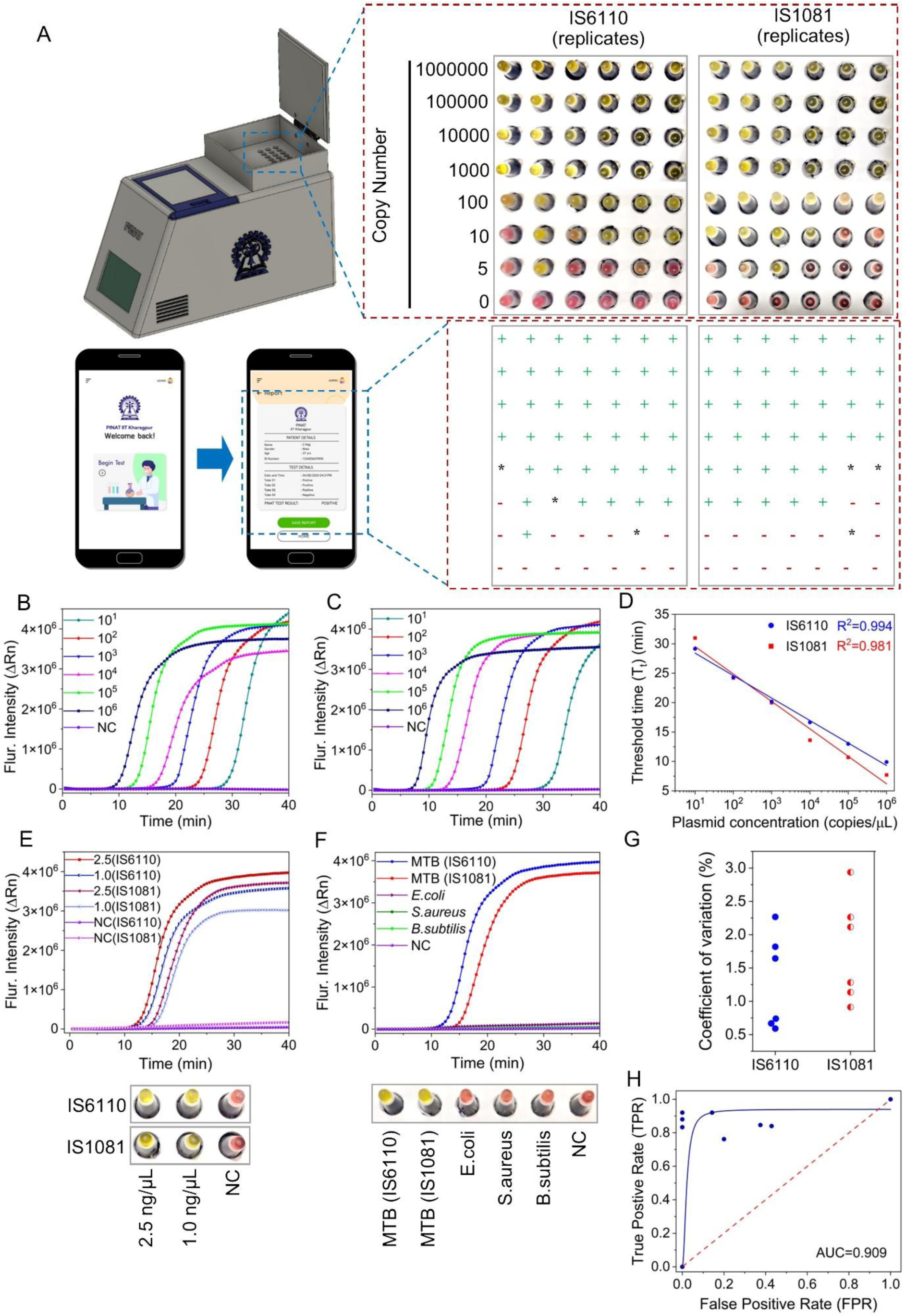
(A) Limit of detection (LoD) of colorimetric screening method for detecting active TB infection or disease targeting two individual genes (IS6110 and IS1081). A known number of copies (ten-fold serial dilutions) of plasmid containing the target gene were amplified through colorimetric LAMP in portable device and subsequently images of tube were captured by integrated camera and finally analyzed by smartphone application. Yellow color corresponds to ‘+’ symbol which denotes positive results, pink color corresponds to ‘-’ symbol which denotes negative results and orange color corresponds to ‘*’ symbol which denotes uninterpreted results. (B) IS6110 and (C) IS1081 targeting limit of detection (LoD) of real-time fluorometric screening method for detecting TB infection/disease. A known number of copies (ten-fold serial dilutions) of plasmid containing the target gene were amplified and detected by real-time fluorometric LAMP. NC: negative control (nuclease-free water). (D) Co-relation between different concentration (copies/µL) of plasmid vs threshold time (min) of real-time LAMP method targeting IS6110 (blue line) and IS1081 (red line). The data was fitted with linear regression (R^2^ =0.994 for IS6110; R^2^ =0.981 for IS1081). (E) Limit of detection (LoD) of the real-time fluorometric and colorimetric detection method targeting both IS6110 and IS1081 using genomic DNA. A known concentrations of genomic DNA of M. tuberculosis were amplified and detected. Real-time amplification curves and yellow color in colorimetric output showing the detection of 2.5 ng/µL and 1 ng/µL of genomic DNA. NC: negative control (nuclease-free water). (F) Specificity of the real-time fluorometric and colorimetric detection method towards the detection different microbial strains. A known concentration of genomic DNA (2.5 ng/µL) from each strain were taken to performed the test. NC: negative control (nuclease-free water). (G) Real-time LAMP reaction was performed five times separately using different copy number of plasmids containing IS6110 and IS1081 gene. Assay precision for both gene was determined by calculating the coefficient of variation between Tt values (threshold time) observed. (H) Receiver operating characteristics (ROC) for the in-vitro assay. Blue solid line indicates the test and the 45° diagonal line (red dotted line) serves as the reference line.

The assay precision in terms of mean coefficient of variation (CV) were found to be within 3% (Fig. 2G, Table S3 and Table S4) which is well below the recommended value of 15% for imprecision [35] thereby indicating exceptional repeatability and reproducibility of the test method. These results altogether demonstrate high sensitivity, specificity and robustness of the test methods for the highly precise detection of the MTB genomic DNA.

The Receiver Operating Characteristic (ROC) curve (Fig. 2H) was plotted to further analyze the test performance on the basis of different concentrations of specific target IS elements inserted in plasmid. The ROC demonstrates the diagnostic ability of a binary classifier system as its discrimination threshold is varied, displaying the trade-off between sensitivity (true positive rate) and specificity (true negative rate) across different threshold values. A higher area under the curve (AUC) denotes better overall performance of the classifier in differentiating between positive and negative cases. Generally, an AUC value of 0.5 indicates a lack of discrimination (meaning the test does not effectively distinguish between patients with and without the disease or condition). An AUC between 0.7 and 0.8 considers acceptable, while 0.8 to 0.9 is classified as excellent, and anything above 0.9 is considered outstanding. The curve was positioned in close proximity to the upper-left corner of the window which ensured that the AUC was 0.909 (95% CI: 0.871–0.947) representing highly favorable trade-off between the sensitivity and specificity at various decision thresholds [36,37].

### Allele specific LAMP for the detection of drug resistant TB infection

Patients infected with more than one strain of MTB other than the WT strain may exhibit phenotypic resistance towards the treatment. If confirmed, patients will receive an alternate regimen of antibiotics. A timely and precise diagnosis of drug resistant TB could help to determine the best course of treatment, lowering rates of morbidity and mortality. We developed a novel and rapid nucleic acid based assay to detect drug resistant TB, which can be easily used for routine diagnosis at resource-limited settings. For this purpose, we have designed a realistic workflow (Fig.S4) for the futuristic implementation of allele specific assay at PHC-level testing environments. Under this approach, TB-LAMP assay will be performed as the primary screening test and resistance assays will be performed only for the samples confirmed as MTB-positive.

Over 95% of mutations that confer resistance to one of the first line drug Rifampicin, reside in the rpoB gene encoding the RNA polymerase β subunit. These mutations cluster within an 81 bp hot spot region known as RIF-resistance determining region (RRDR) that harbours codons 426 to 452 (MTB nomenclature). The highest global incidence of RIF resistances is linked to the group I mutations at codons 435, 445, and 450. In contrast, majority (60-80%) of the isoniazid resistance originate from a single point mutation at the codon 315 in the katG gene [38,39]. Based upon the prevalence of the drug resistance mutations associated with the MDR-TB cases reported in India [2] we targeted four major mutations, rpoB D435V, rpoB H445D, S450L, and katG S315T, associated with rifampicin- and isoniazid-resistant TB infections respectively.

Allele specific PCR utilizes a single nucleotide mismatch in the 3’ penultimate nucleotide of the primer in order to detect and distinguish single nucleotide polymorphism (SNP) in mutational alleles [40–42]. We improvised this methodology to develop allele specific LAMP, where the single nucleotide mismatch was introduced in the 3’ penultimate nucleotide of the FIP. The 3’ terminal nucleotide kept complementary to the mutational allele so that for the WT allele the FIP will encounter two consecutive mismatches at the 3’-end. This should increase the sensitivity of the mutational alleles several folds over the WT thereby result in the selective amplification of the former over the later. In order to verify the feasibility of our proposed principle (Fig. 3A) and to demonstrate the potential of the allele specific LAMP to detect MDR-TB specific mutations, we designed different combination of allele-specific primers and evaluated their efficacy in differentiating WT and MT alleles. These primers were empirically screened and qualitatively scored based on their ability to effectively amplify the MT over the WT alleles under the isothermal condition (Table 1). As evidenced (Fig. 3 B-E), FIPs that carried no base modification at penultimate position (−1 position adjacent to SNP 3’ end), showed comparable amplification of the MT and the WT rpoB and katG templates in both colorimetric and real-time fluorometric assay. In contrast, when FIPs with at least 1 nucleotide mismatches at −1 position of SNP at 3’end was used in the reaction, the difference in amplification speed (threshold time Tt) between the WT and MT templates was remarkable. The mutant experienced 1 mismatch whereas WT experienced 2 mismatches during amplification. The discrimination between WT and MT of rpoB H445D and katG S315T were successfully achieved depending on this strategy. No amplification was observed for the WT hence no color change was visualized. In contrast the MT alleles showed distinct color change within stipulated time period. In real time LAMP, the difference in threshold time between WT versus MT alleles became greater than 40 min (ΔTt> 40 min) (Fig. 3C and 3E) thus achieving high efficiency detection of the above-mentioned mutations.

**Fig. 3.**
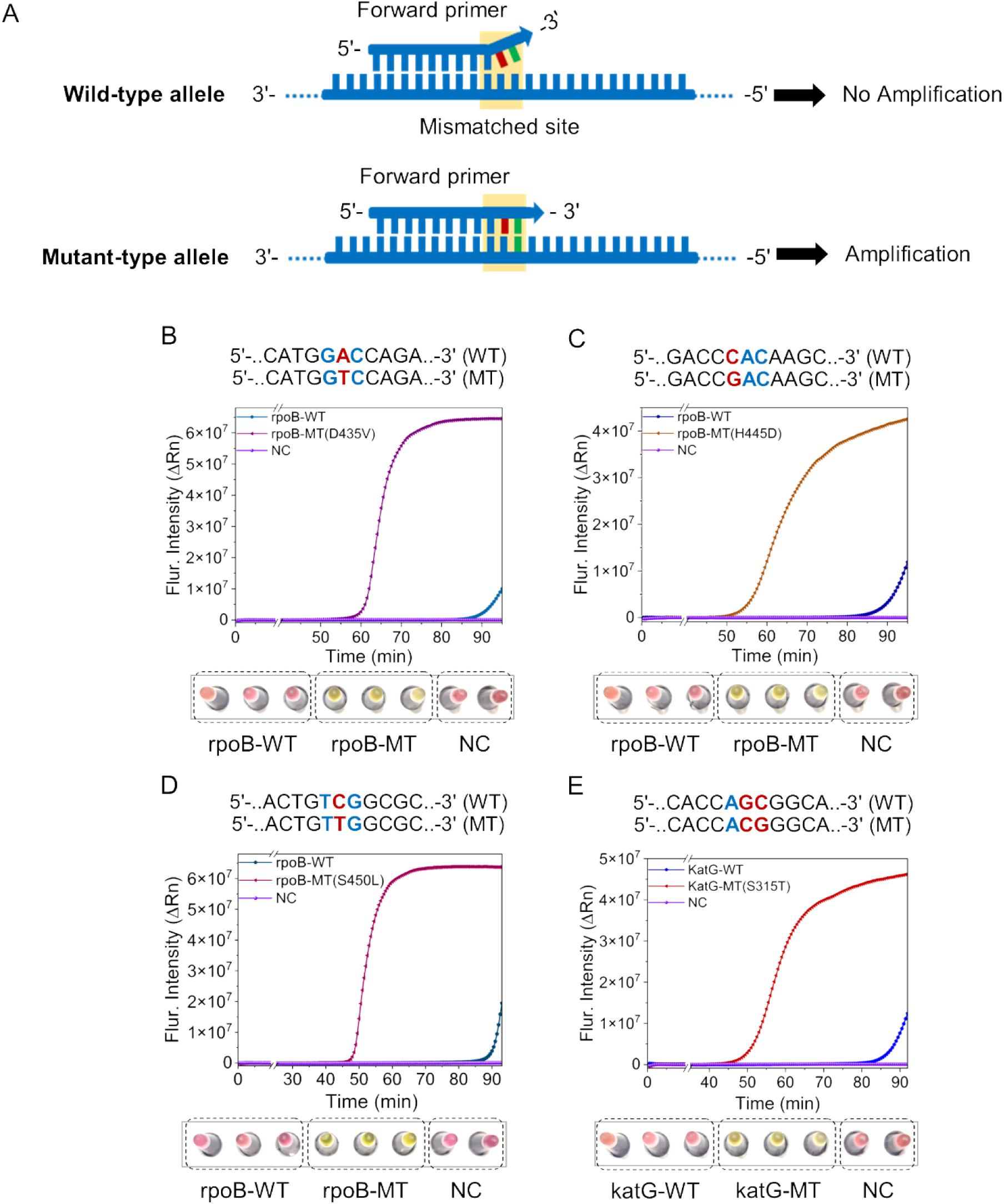
Allele specific LAMP for the detection of rifampicin and isoniazid resistant MTB. (A) Allele specific primers were designed to help better discriminate the MT alleles from WT alleles. The forward primer (F2 region of FIP) was kept fully complimentary to the MT sequences whereas it was partially complimentary to the WT sequence. To detect MT alleles, one or two mismatches was introduced at penultimate position (−1, or −1 and −2) adjacent to SNP binding site of the 3′ end of the primer. During LAMP reaction, the primer with two or three mismatches towards WT DNA, inhibited annealing of the primer; thus, suppressing the amplification of WT DNA. Real-time amplification plot showing the amplification of MT DNA having point mutation at (B) rpoB D435V, (C) rpoB H445D, (D) rpoB S450L, and (E) katG S315T site. MT DNA showed exponential curve in each case. Corresponding colorimetric results have been showed below the real-time plot. Reactions were performed in triplicate. Yellow color was visible for successful reaction. WT: Wild type, MT: Mutant type, NC: Negative control (nuclease-free water).

**Table 1.**
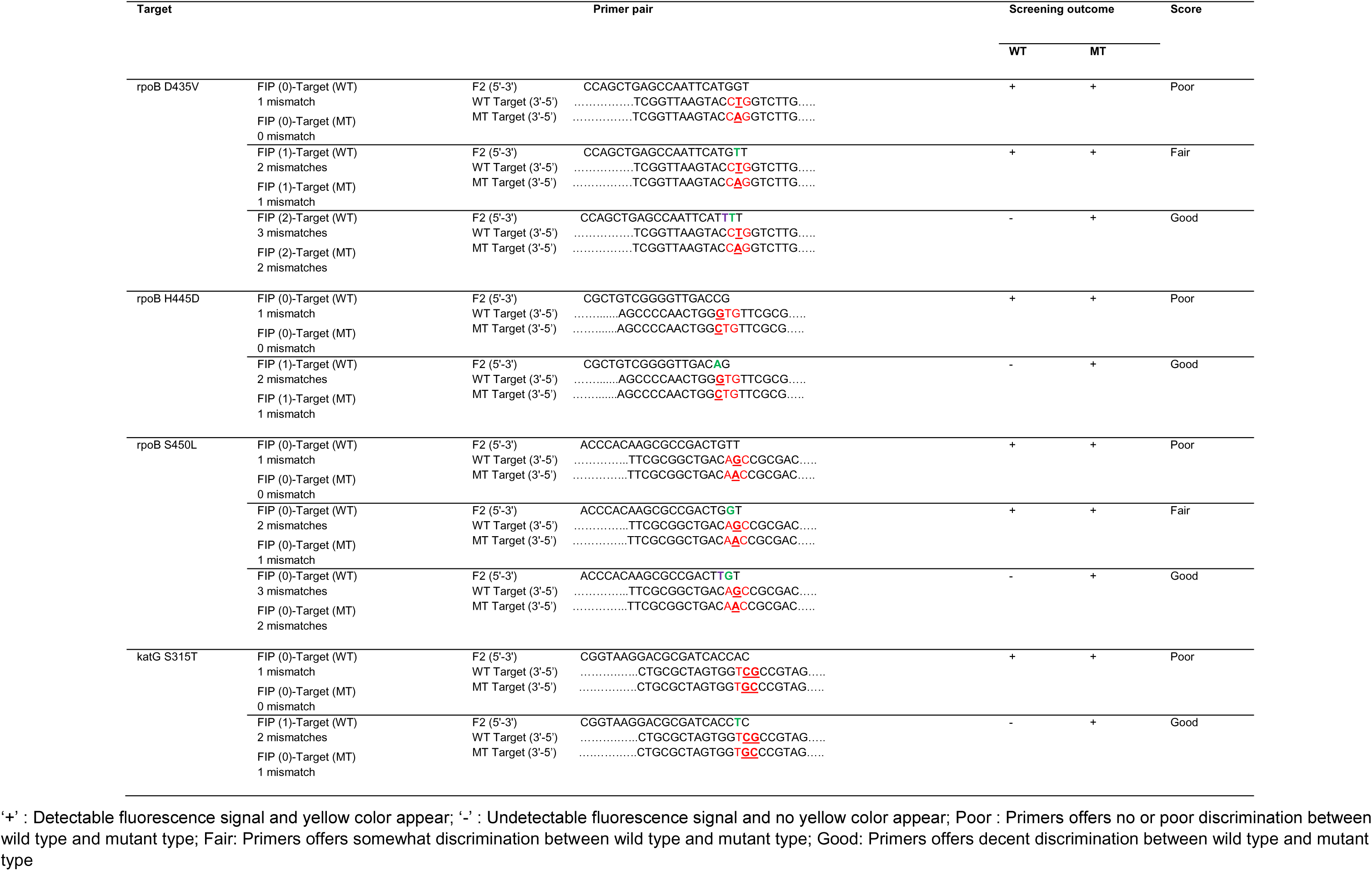
Preliminary screening results of FIP primer pairs carrying 0–3 base mismatches in their sequences. The primer pairs were scored according to their ability to manoeuvre the LAMP amplification toward the target based on the fluorescence signals and colorimetric outcome between mutant and wild-type allele.

Interestingly for the rpoB D435V and S450L mutations, a single mismatch at the 3’ penultimate position could not discriminate the WT versus MT alleles. To address this issue, a consecutive two nucleotide mismatch was introduced upstream (−1 and −2 position) to SNP site from the 3’ end of the allele-specific FIP. Herein, the MT allele experienced 2 mismatches whereas WT allele experienced 3 mismatches during the annealing with the allele specific primer. Notably, this FIP (Fig 3B and 3D) significantly supressed the WT amplification compared to the MT as evidenced for both real-time fluorescence (ΔTt> 40 min) and colorimetric reactions. Thus, adjustment of LAMP-FIP primers through incorporation of base mismatches at the 3’-end is critical for preferential amplification of the SNP target by altering the free energy for binding between the primer with the MT versus WT alleles. This preferential amplification of the MT versus WT alleles is achieved without using any additional molecular probes, or analyses and hence has a remarkable clinical implication in terms of detection of drug resistant TB infections in resource limited settings.

### Efficacy evaluation of TB-LAMP assay for detecting TB infections in clinical samples

The IS6110 and IS1081 targeting TB-LAMP assay showed high sensitivity and specificity in the laboratory based *in-vitro* standardization experiments. So, we aimed to assess the clinical performance of this assay using extracted DNA samples from the sputum. To ensure high analytical specificity and mitigate potential sources of error, a dual-target gene testing strategy was employed at ICMR-NIRT, Chennai (Fig. 4A). Previously reported LAMP based MTB detection methods [11,43,44] relying solely upon IS6110 showed variable 50-90% sensitivity and 60-100% specificity. A few LAMP-based studies targeting IS1081 showed varied range of sensitivity (25%-78%) and specificity (80-100%) [45–47]. In our study, within the smartphone application-enabled detection algorithm, samples were classified as positive or negative only when both target genes provided concordant positive or negative result, respectively. Samples in which only one of the two target genes tested positive were classified as presumptive positive and subjected to repeat testing. If the repeat assay reproduced the same gene-specific positivity, the sample was interpreted as TB positive in accordance with CBNAAT (GeneXpert) results; otherwise, it was reported as negative. However, we have demonstrated gene wise sensitivity and specificity. A positive signal for the internal control (RNase P) in a sample that was negative or positive for the MTB target (IS6110 and IS1081) confirms the test was valid and the sample was indeed negative or positive, based on the outcome of MTB target genes. Otherwise, the test has been considered as invalid. Successful amplification of the ‘positive amplification control’ confirmed the integrity and performance of each batch of the reaction kit. In addition, development of orange coloration after amplification failed to provide smartphone application-based interpretation. Inconsistent amplification of internal control or positive amplification control irrespective of MTB target amplification, or the developing of orange coloration after reaction, was regarded as an ‘uninterpreted result’. In this study, the proportion of uninterpretable result was within the accepted range (<4%) [20,48] The ‘uninterpreted’ results were excluded from the overall performance calculations.

**Fig. 4.**
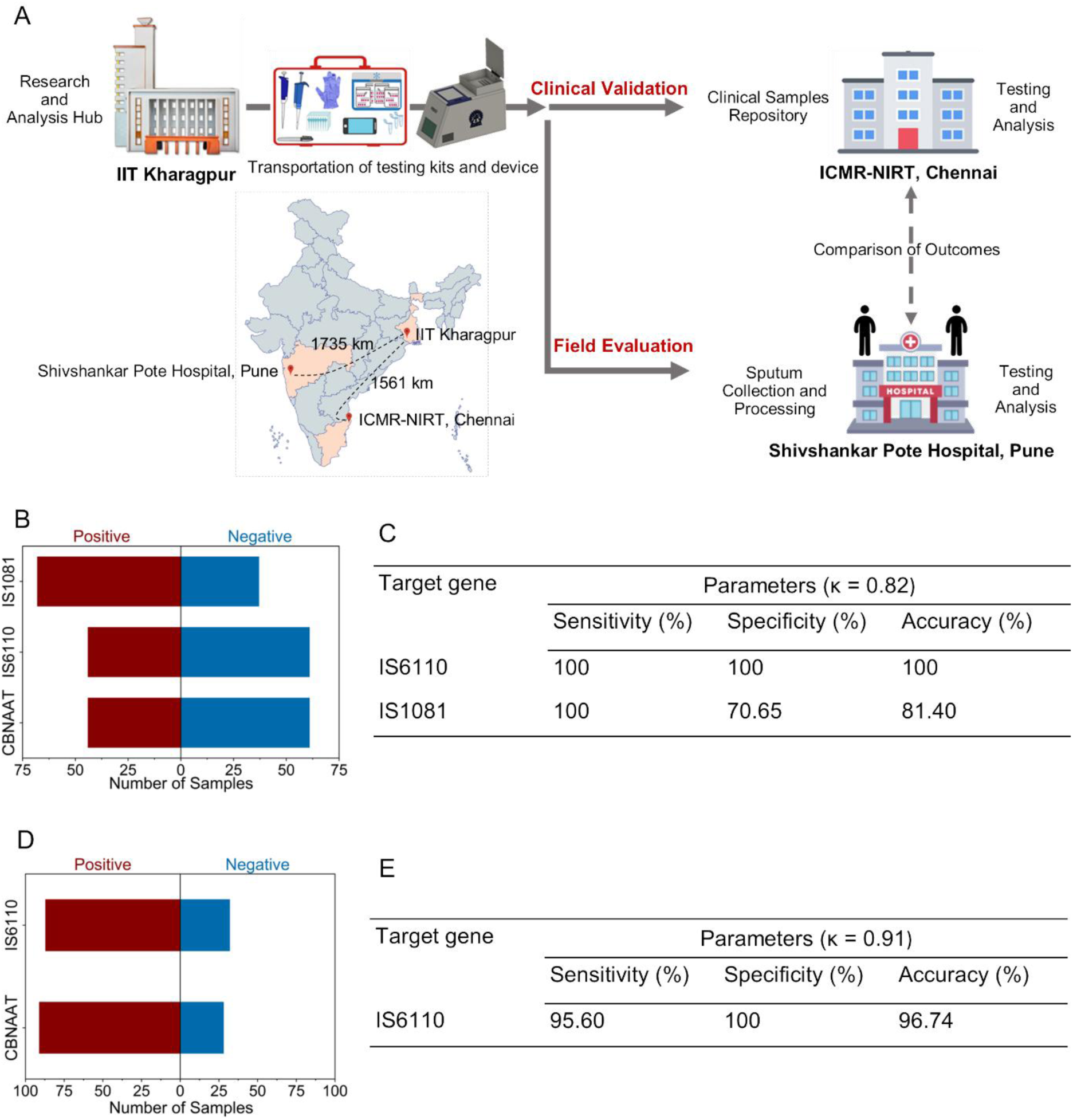
(A) Workflow for clinical validation and field evaluation of colorimetric TB screening test at ICMR-NIRT, Chennai (laboratory set-up) and Shivshankar Pote Hospital, Pune (resource-limited settings). The location and distance of the sites have been shown in the map. The testing kits and POC devices were manufactured at IIT Kharagpur from where the test kits and the devices were transported to the respective centres in order to detect TB infections/disease. In field evaluation, sample processing, and detection process were performed at the centre. All test results were analysed and compared with CBNAAT (GeneXpert) and upload to the database at analysis hub. (B) Comparative distribution of positive and negative population of TB infections against CBNAAT (GeneXpert), obtained by testing 105 genomic DNA, isolated from human sputum samples during clinical validation (C) Gene wise comparative clinical sensitivity, specificity and accuracy of two-gene test for TB infection detection. (D) Comparative distribution of positive and negative population of TB infections against CBNAAT (GeneXpert), obtained by testing 119 genomic DNA, isolated from human sputum samples during field evaluation of the test. (E) Clinical sensitivity, specificity and accuracy of single gene test for TB infection detection.

Our method showed considerably high sensitivity, specificity and accuracy for screening clinical samples, when compared against the benchmark CBNAAT (GeneXpert) test (Fig.4B, 4C). The assay targeting IS6110 demonstrated a sensitivity and specificity of 100% (95% CI: 97.14% - 100%) whereas for the IS1081, the sensitivity and specificity were 100% (95% CI: 97.14% - 100%) and 70.65% (95% CI: 59.06% - 81.94%) respectively. In addition, the assay also demonstrated 100% accuracy for the IS6110 target and an accuracy of 81.4% for the IS1081 target. When both targets were considered together, the overall accuracy of the two-gene assay exceeded 90%. The IS6110 which showed remarkable sensitivity, specificity and gene specific accuracy in our assay, may be considered as preferred target for detecting TB infection and disease as it is noticed in majority of the MTB strains isolated from Indian population. *M. tuberculosis* H37Rv contains 16 copies of IS6110 and 5-6 copies of IS1081 thereby further confirming the suitability of the IS6110 as a robust biomarker for MTB screening. Nonetheless, combining IS6110 and IS1081 would further increase the durability of the test procedure when deployed for large population in community level screening. We also confirmed this fact, the level of reliability and concordance with the benchmark test by analysing Cohen’s kappa (k) statistics. The combined average κ value (κ = 0.82) of the two-gene assay demonstrated as ‘near-perfect agreement’ with the benchmark test and was also consistent with previously reported thresholds [27,28,49], and indicates a high level of reliability of the assay as a molecular diagnostic test (Fig. 4C). Together, the TB-LAMP assay showed high efficacy for the detection of active TB infection or disease with clinical samples in laboratory environment.

### Evaluation of a single gene TB-LAMP assay for the detection of TB infections at resource-limited settings

Till date, as per our knowledge, the availability of LAMP-based assay platform integrated with fully portable device and smartphone app-based data interpretation capable of detecting TB at the point of sample collection with minimal infrastructure, remains limited [20]. So, encouraged by the remarkable sensitivity and specificity of the IS6110 targeting LAMP assay during the preliminary validation with clinical samples, we initially aimed to deploy a single gene colorimetric TB-LAMP assay for the detection of TB infection or disease directly from live patient samples in a primary healthcare (PHC) setting (Fig. 4A). Single-gene diagnostic assays may be significantly reduced per-test cost and logistical complexity, facilitating large-scale field implementation by personnel with minimal technical training. However, reliance on a single genetic target carries an inherent risk of false-negative or false-positive results, which can be mitigated through multi-gene testing strategies. But, herein, we initially tried to assess the practicality and performance of our proposed strategy and technology in real-world field settings. Accordingly, after obtaining ethical and relevant regulatory approvals, the one gene colorimetric TB-LAMP assay was deployed at Shivshankar Pote Hospital, Pune Municipal Corporation, India. This healthcare centre has minimal laboratory facility mimicking the perfect scenario of resource-limited setting of PHC. Although, testing kits were transported from a long distance of IIT Kharagpur, the stability of testing kits remained unaltered. To confirmed it, we investigated the stability and QC of each batch of testing kit. Testing kit storage at various temperature conditions for one month (Fig. S5) revealed high stability of the kit components at −20^0^C and at 4^0^C conditions, even after the long-distance transport. The single gene test set-up and validation of test based on the outcome of internal control and validation kit batch based on the outcome of positive amplification control were followed as defined during earlier clinical validation at ICMR-NIRT, Chennai. Samples in which single gene tested positive, were classified as positive, otherwise it was reported as negative. The proportion of uninterpretable result under predefined criteria, was within the accepted range (<4%) [20,48]. The ‘uninterpretable’ results were excluded from the overall performance calculations.

Herein, the TB-LAMP assay showed high sensitivity, specificity and accuracy level for detecting active TB infection or disease based on the IS6110, when compared against the benchmark CBNAAT test (GeneXpert). The sensitivity and specificity of the test were found as 95.60% (95% CI: 91.39% - 99.81%) and 100% (95% CI: 97.48 −100%) respectively (Fig.4D, 4E). Additionally, the single-gene assay demonstrated an overall accuracy of approximately 97%. The single-gene assay demonstrated near-perfect agreement with the benchmark test, as reflected by Cohen’s kappa value (κ = 0.91), indicating a high level of assay reliability (Fig.4E). The overall outcome with cohort details have been provided in Table S5.

The initial clinical evaluation and this field exercise altogether confirmed that this TB-LAMP assay integrated with this portable device makes this testing ideal to be implemented at the resource-limited settings without compromising the detection accuracy and reliability. In addition, we investigated that the cost per test is also much lower than the existing common molecular test, which will provide additional advantage over the available predicate technologies. A brief comparison has been provided based on the cost per test, time, easiness and other important parameters in Table S6.

As a limitation, we believe that an in-house, rapid DNA extraction procedure need to be incorporated in the protocol for a comprehensive field-ready package. We also acknowledge that the samples were obtained from a limited number (two) of sites, and therefore may not fully capture the geographic and epidemiological diversity of the broader TB patient population across India because evaluating the broader TB patient population in a large country like India, is always be highly challenging in a short period. Nonetheless, these attempts appear to be a significant milestone towards bringing low-cost high-end molecular diagnostics from sophisticated labs to under-resourced community settings without unacceptable compromise in the predictive accuracy.

## CONCLUSIONS

Through this study, we have introduced a low-cost, user-friendly, and rapid point-of-care diagnostic platform for tuberculosis, built on a dual-marker LAMP assay and integrated into a portable device synchronized with a smartphone application. The system demonstrated excellent analytical and clinical performance with achieving exceptional sensitivity and specificity in laboratory-based evaluation as well as in resource-limited settings. Beyond the initial TB screening, we have also developed an allele-specific LAMP-based assay capable of identifying key mutations originating from the resistance of widely used first-line drugs, i.e., rifampicin and isoniazid, offering a pathway towards rapid and accessible diagnosis of MDR-TB. Together, these advances mark a significant step toward democratizing TB diagnostics. By combining robust molecular assays with portable, AI-enabled technology, our platform addresses longstanding barriers to early detection and treatment in high-burden regions. With further validation, this approach holds promise to transform community-level screening, curb MDR-TB transmission, and accelerate progress towards the global TB elimination goals.

## AUTHOR CONTRIBUTIONS

**Sudip Nag:** Writing-original draft, Writing-review & editing, Conceptualization, Visualization, Validation, Methodology, Investigation, Data curation, Formal analysis. **Saptarshi Banerjee:** Formal analysis, Data curation (clinical validation at ICMR-National Institute for Research in Tuberculosis, Chennai), **Sohom Banerjee:** Data curation (smartphone application). **Subham Ghosh:** Data curation (field validation at Shivshankar Pote Hospital, Pune). **Arijit Bera:** Data curation (allele-specific assay), **Sivakumar Shanmugam:** Clinical validation. **Arindam Mondal:** Writing-review & editing, Supervision, Conceptualization, Resources, Project administration, Funding acquisition. **Suman Chakraborty:** Writing-review & editing, Supervision, Conceptualization, Resources, Project administration, Funding acquisition

## FINANCIAL SUPPORT AND ACKNOWLEDGEMENTS

All authors acknowledge the Indian Institute of Technology Kharagpur, and the Indian Council for Medical Research (ICMR) for facilitating different components of the work. Further, the authors profoundly acknowledge ICMR-National Institute for Research in Tuberculosis, Chennai, India for supporting the clinical validation and Shivshankar Pote Hospital, Pune, India for supporting field evaluation of the test. S.C. acknowledges SERB, Department of Science and Technology, Government of India, for their support through Sir J. C. Bose National Fellowship. A.M. also acknowledges ICMR funded Extramural CAR project [EM/DEV/CAR/2/0517/2023 (EOFFICE-179944), Dt:18.04.2024] and the Department of Science and Technology & Biotechnology (DSTBT), Government of West Bengal (GoWB) funded project [1111(Sanc.)/STBT-11012(12)/2/2024-WBSCST SEC] for the partial financial support to carry out this research work. S.C and A.M also profoundly acknowledge ICMR-DHR for their financial support under the Center of Excellence Program on Healthcare. SC acknowledges DSIR, Ministry of Science and Technology, Government of India for their financial support under the Common Research and Technology Development Hub (CRTDH) on Affordable Healthcare at IIT Kharagpur. S.N. acknowledges ICMR for providing Research Associate fellowship [(i). File No: 5/3/8/14/ITR-F/2022-ITR, Dt. 11.02.2022 and (ii). Sanc. No: IIT SRIC IDK PHASE III 2024, Dt. 27.11.2024]. The authors acknowledge Mr. Akasdeep Mondal for his support in device design, fabrication and Mr. Rana Akhter for helping with the smartphone application development. The Authors also acknowledge Mr. Sambit Ghosh and Mr. Rohit Bajaj for their support during the field evaluation of the test at Shivshankar Pote Hospital, Pune, India.

## Supporting information

Supplementary Informations

## Data Availability

The data supporting the findings of this study are available within the manuscript and its supplementary information. The raw and analyzed datasets generated during the study are available for research purposes from the corresponding authors on reasonable request.

## DISCLAIMER

The funding sources had no influence on the study design, data collection, analysis, interpretation of the data, or the writing of the manuscript. The content of this work is solely the responsibility of the authors.

## ETHICS APPROVAL

This study has been approved by Indian Institute of Technology Kharagpur Biosafety and Ethics Committee obeying ethical principles for medical research involving human subjects according to the declaration of Helsinki guideline and obeying the guidelines of the Indian Council for Medical Research (ICMR). In addition, for the investigation involving human subjects, informed consent has been obtained from the participant involved.

## REFERENCES

1. Global tuberculosis report 2025. Geneva: World Health Organization; 2025. Licence: CC BY-NC-SA 3.0 IGO.

2. Indian Catalogue of *Mycobacterium tuberculosis* Mutations and their Association with Drug Resistance Version 2.0. 2024. ICMR-National Institute for Research in Tuberculosis.

3. Catalogue of mutations in Mycobacterium tuberculosis complex and their association with drug resistance, second edition. Geneva: World Health Organization; 2023. Licence: CC BY-NC-SA 3.0 IGO.

4. Yusoof KA, García JI, Schami A, Garcia-Vilanova A, Kelley HV, Wang S-H, Rendon A, Restrepo BI, Yotebieng M, Torrelles JB. 2022. Tuberculosis Phenotypic and Genotypic Drug Susceptibility Testing and Immunodiagnostics: A Review. Front Immunol 13:870768.

5. Siddiqi S, Ahmed A, Asif S, Behera D, Javaid M, Jani J, Jyoti A, Mahatre R, Mahto D, Richter E, Rodrigues C, Visalakshi P, Rüsch-Gerdes S. 2012. Direct Drug Susceptibility Testing of Mycobacterium tuberculosis for Rapid Detection of Multidrug Resistance Using the Bactec MGIT 960 System: a Multicenter Study. J Clin Microbiol 50:435–440.

6. MacLean E, Kohli M, Weber SF, Suresh A, Schumacher SG, Denkinger CM, Pai M. 2020. Advances in Molecular Diagnosis of Tuberculosis. J Clin Microbiol 58:e01582–19.

7. Chakravorty S, Simmons AM, Rowneki M, Parmar H, Cao Y, Ryan J, Banada PP, Deshpande S, Shenai S, Gall A, Glass J, Krieswirth B, Schumacher SG, Nabeta P, Tukvadze N, Rodrigues C, Skrahina A, Tagliani E, Cirillo DM, Davidow A, Denkinger CM, Persing D, Kwiatkowski R, Jones M, Alland D. 2017. The New Xpert MTB/RIF Ultra: Improving Detection of *Mycobacterium tuberculosis* and Resistance to Rifampin in an Assay Suitable for Point-of-Care Testing. mBio 8:e00812–17.

8. McNerney R, Daley P. 2011. Towards a point-of-care test for active tuberculosis: obstacles and opportunities. Nat Rev Microbiol 9:204–213.

9. Boehme CC, Nicol MP, Nabeta P, Michael JS, Gotuzzo E, Tahirli R, Gler MT, Blakemore R, Worodria W, Gray C, Huang L, Caceres T, Mehdiyev R, Raymond L, Whitelaw A, Sagadevan K, Alexander H, Albert H, Cobelens F, Cox H, Alland D, Perkins MD. 2011. Feasibility, diagnostic accuracy, and effectiveness of decentralised use of the Xpert MTB/RIF test for diagnosis of tuberculosis and multidrug resistance: a multicentre implementation study. The Lancet 377:1495–1505.

10. Lin M, Chen Y-W, Li Y-R, Long L-J, Qi L-Y, Cui T-T, Wu S-Y, Lin J-Y, Wu T, Yang Y-C, Yuan W-H, Wu G-Y, Lan Q-W, Liu J-Q, Li Y-P, Yu Z-Y, Guo X-G. 2022. Systematic evaluation of line probe assays for the diagnosis of tuberculosis and drug-resistant tuberculosis. Clinica Chimica Acta 533:183–218.

11. Araújo RM, Montenegro RDA, Peixoto ADS, Silva LLDS, Da Costa RMPJ, Carvalho-Silva WHV, Mello DCD, Diniz GTN, Silva MALD, Melo FLD, Montenegro LML, Schindler HC. 2023. Performance of IS6110-LAMP assay for detection of Mycobacterium tuberculosis complex in blood and urine samples from patients with extrapulmonary tuberculosis. Tuberculosis 143:102423.

12. Batuer M, Yuan Y, Yu M, Meng C. 2024. Establishment and evaluation of a new fluorescent probe method based on loop-mediated isothermal amplification for the detection of *Mycobacterium tuberculosis* complex. Luminescence 39:e4795.

13. Nag S, Banerjee S, Bandopadhyay A, Banerjee I, Jana S, Mondal A, Chakraborty S. 2025. CO-INFECTS: A highly affordable, portable, nucleic acid-based rapid detector for differential diagnostics of active respiratory co-infections. Sensors and Actuators B: Chemical 433:137516.

14. Tomita N, Mori Y, Kanda H, Notomi T. 2008. Loop-mediated isothermal amplification (LAMP) of gene sequences and simple visual detection of products. Nat Protoc 3:877–882.

15. Mori Y, Notomi T. 2009. Loop-mediated isothermal amplification (LAMP): a rapid, accurate, and cost-effective diagnostic method for infectious diseases. Journal of Infection and Chemotherapy 15:62–69.

16. Gray CM, Katamba A, Narang P, Giraldo J, Zamudio C, Joloba M, Narang R, Paramasivan CN, Hillemann D, Nabeta P, Amisano D, Alland D, Cobelens F, Boehme CC. 2016. Feasibility and Operational Performance of Tuberculosis Detection by Loop-Mediated Isothermal Amplification Platform in Decentralized Settings: Results from a Multicenter Study. J Clin Microbiol 54:1984–1991.

17. Boehme CC, Nabeta P, Henostroza G, Raqib R, Rahim Z, Gerhardt M, Sanga E, Hoelscher M, Notomi T, Hase T, Perkins MD. 2007. Operational Feasibility of Using Loop-Mediated Isothermal Amplification for Diagnosis of Pulmonary Tuberculosis in Microscopy Centers of Developing Countries. J Clin Microbiol 45:1936–1940.

18. Schijman AG, Alonso-Padilla J, Britto C, Herrera Bernal CP. 2024. Retrospect, advances and challenges in Chagas disease diagnosis: a comprehensive review. The Lancet Regional Health - Americas 36:100821.

19. Jin A, Deng M, Yang HS, Li Z. 2026. Loop-mediated isothermal amplification (LAMP)-based microbial detection: a review of FDA-authorized tests and future perspectives. Critical Reviews in Clinical Laboratory Sciences 63:57–79.

20. The Use of Loop-Mediated Isothermal Amplification (TB-LAMP) for the Diagnosis of Pulmonary Tuberculosis: Policy Guidance. World Health Organization, Geneva. http://www.ncbi.nlm.nih.gov/books/NBK384520/. Retrieved 6 April 2026.

21. Zaber M, Hoque F, Paean IM, Tarafder S. 2024. Evaluation of Multiplex loop-mediated isothermal amplification assay for the detection of Mycobacterium tuberculosis complex from clinically suspected cases of pulmonary tuberculosis. Heliyon 10:e39847..

22. Jirakittiwut N, Ratthawongjirakul P. 2025. Advances in Isothermal Amplification for the Diagnosis of Tuberculosis. Clinical Laboratory Analysis 39:e70113.

23. Sun W, Li C, Wu X, Li Y, Yang L, Liu Y, Cheng X, Guo S, Ma L, Qiu H, Du W. 2025. Point-of-care detection of Mycobacterium in bovine feces using a portable real-time loop-mediated isothermal amplification system. Sensors and Actuators B: Chemical 440:137857.

24. Ying G-S, Maguire MG, Glynn RJ, Rosner B. 2020. Calculating Sensitivity, Specificity, and Predictive Values for Correlated Eye Data. Invest Ophthalmol Vis Sci 61:29.

25. Hanley JA. 1983. If Nothing Goes Wrong, Is Everything All Right?: Interpreting Zero Numerators. JAMA 249:1743.

26. Eypasch E, Lefering R, Kum CK, Troidl H. 1995. Probability of adverse events that have not yet occurred: a statistical reminder. BMJ 311:619–620.

27. McHugh ML. 2012. Interrater reliability: the kappa statistic. Biochem Med (Zagreb) 22:276–282.

28. Li M, Gao Q, Yu T. 2023. Kappa statistic considerations in evaluating inter-rater reliability between two raters: which, when and context matters. BMC Cancer 23:799.

29. Fomukong N, Beggs M, El Hajj H, Templeton G, Eisenach K, Cave MD. 1998. Differences in the prevalence of IS6110 insertion sites in Mycobacterium tuberculosis strains: low and high copy number of IS6110. Tubercle and Lung Disease 78:109–116.

30. Kremer K, Glynn JR, Lillebaek T, Niemann S, Kurepina NE, Kreiswirth BN, Bifani PJ, Van Soolingen D. 2004. Definition of the Beijing/W Lineage of *Mycobacterium tuberculosis* on the Basis of Genetic Markers. J Clin Microbiol 42:4040–4049.

31. Nagamine K, Hase T, Notomi T. 2002. Accelerated reaction by loop-mediated isothermal amplification using loop primers. Molecular and Cellular Probes 16:223–229.

32. Liang L, Chen M, Hu O, He Q, Chen Z. 2023. A novel fluorescence method based on loop-mediated isothermal amplification and universal molecular beacon in Mycobacterium tuberculosis detection. Talanta 253:123996.

33. Wen J, Gou H, Wang S, Lin Q, Chen K, Wu Y, Huang X, Shen H, Qu X, Lin J, Liao M, Zhang J. 2021. Competitive activation cross amplification combined with smartphone-based quantification for point-of-care detection of single nucleotide polymorphism. Biosensors and Bioelectronics 183:113200.

34. Jaroenram W, Kampeera J, Arunrut N, Karuwan C, Sappat A, Khumwan P, Jaitrong S, Boonnak K, Prammananan T, Chaiprasert A, Tuantranont A, Kiatpathomchai W. 2020. Graphene-based electrochemical genosensor incorporated loop-mediated isothermal amplification for rapid on-site detection of Mycobacterium tuberculosis. Journal of Pharmaceutical and Biomedical Analysis 186:113333.

35. Smith G. 2010. Bioanalytical Method Validation: Notable Points in The 2009 Draft EMA Guideline and Differences with The 2001 FDA Guidance. Bioanalysis 2:929–935.

36. Nahm FS. 2022. Receiver operating characteristic curve: overview and practical use for clinicians. Korean J Anesthesiol 75:25–36.

37. Hajian-Tilaki K. 2013. Receiver Operating Characteristic (ROC) Curve Analysis for Medical Diagnostic Test Evaluation. Caspian J Intern Med 4:627–635.

38. Zaw MT, Emran NA, Lin Z. 2018. Mutations inside rifampicin-resistance determining region of rpoB gene associated with rifampicin-resistance in Mycobacterium tuberculosis. Journal of Infection and Public Health 11:605–610.

39. Singpanomchai N, Akeda Y, Tomono K, Tamaru A, Santanirand P, Ratthawongjirakul P. 2021. Rapid detection of multidrug-resistant tuberculosis based on allele-specific recombinase polymerase amplification and colorimetric detection. PLoS ONE 16:e0253235.

40. Suzuki Y, Sekiya T, Hayashi K. 1991. Allele-specific polymerase chain reaction: A method for amplification and sequence determination of a single component among a mixture of sequence variants. Analytical Biochemistry 192:82–84.

41. Ugozzoli L, Wallace R. 1991. Allele-specific polymerase chain reaction. Methods 2:42–48.

42. Kalendar R, Shustov AV, Akhmetollayev I, Kairov U. 2022. Designing Allele-Specific Competitive-Extension PCR-Based Assays for High-Throughput Genotyping and Gene Characterization. Front Mol Biosci 9:773956.

43. Dale JW, Tang TH, Wall S, Zainuddin ZF, Plikaytis B. 1997. Conservation of IS6110 sequence in strains of Mycobacterium tuberculosis with single and multiple copies. Tubercle and Lung Disease 78:225–227.

44. Sharma M, Sharma K, Sharma A, Gupta N, Rajwanshi A. 2016. Loop-mediated isothermal amplification (LAMP) assay for speedy diagnosis of tubercular lymphadenitis: The multi-targeted 60-minute approach. Tuberculosis 100:114–117.

45. Yang B, Wang X, Li H, Li G, Cao Z, Cheng X. 2011. Comparison of loop-mediated isothermal amplification and real-time PCR for the diagnosis of tuberculous pleurisy: LAMP for the diagnosis of tuberculous pleurisy. Letters in Applied Microbiology 53:525–531.

46. Sharma M, Singh R, Sharma A, Gupta V, Sharma K. 2022. IS1081-based Multi-targeted LAMP: An Opportunity to Detect Tubercular Uveitis. Ocular Immunology and Inflammation 30:168–173.

47. Batuer M, Yuan Y, Yu M, Meng C. 2024. Establishment and evaluation of a new fluorescent probe method based on loop-mediated isothermal amplification for the detection of *Mycobacterium tuberculosis* complex. Luminescence 39:e4795.

48. Boehme CC, Nabeta P, Hillemann D, Nicol MP, Shenai S, Krapp F, Allen J, Tahirli R, Blakemore R, Rustomjee R, Milovic A, Jones M, O’Brien SM, Persing DH, Ruesch-Gerdes S, Gotuzzo E, Rodrigues C, Alland D, Perkins MD. 2010. Rapid Molecular Detection of Tuberculosis and Rifampin Resistance. N Engl J Med 363:1005–1015.

49. Callahan C, Ditelberg S, Dutta S, Littlehale N, Cheng A, Kupczewski K, McVay D, Riedel S, Kirby JE, Arnaout R. 2021. Saliva is Comparable to Nasopharyngeal Swabs for Molecular Detection of SARS-CoV-2. Microbiol Spectr 9:e00162–21.

