## Supplementary Informations for "Field-ready portable rapid nucleic acid test for tuberculosis detection and drug-resistance profiling in resource-limited settings"

### ADDITIONAL METHODS

#### Designing assay primers for detecting active and drug resistant TB infection

For screening active MTB infection, two primer sets were designed against two conserved target regions in the insertion elements IS6110 and IS1081 present in the MTBC genome. Whole genome sequences of the organisms were retrieved from NCBI GenBank (http://www.ncbi.nlm.nih.gov/). Copy number of the IS6110 and IS1081 were determined by nucleotide BLAST (https://blast.ncbi.nlm.nih.gov/Blast.cgi). CIRCOS maps to determine the position and number of IS elements for each organism were generated by Python using Matplotlib. In each organism the sequences of different copies of the IS elements were aligned to determine the possible polymorphism. Nucleotide sequence were aligned using Multiple Alignment Fast Fourier Transform (MAFFT) as a high-speed multiple sequence alignment program. Positional nucleotide summary was analyzed from the multiple alignment file in Bio-edit sequence editor. The conserved nucleotides position was selected as target for the primer designing. Then six sets of primers i.e. forward outer primer (F3), backward outer primer (B3), forward inner primer (FIP), backward inner primer (BIP), loop forward primer (LF) and loop backward primer (LB) were designed on the basis of these sequences using PrimerExplorer V5 software (Fujitsu Limited, Tokyo, Japan).

Additionally, allele specific LAMP assay was developed targeting the most common rifampicin- and isoniazid-associated mutations, which are located at codons 435, 445 & 450 of Rifampicin Resistance Determining Region (RRDR) of rpoB gene and at codon 315 of katG gene, respectively [1-3]. Different primer sets were initially designed with the assistance of PrimerExplorer V5 from the reference DNA sequence of *M. tuberculosis* H37Rv (GenBank accession no. AL123456.3) and subsequently the forward inner primer (FIP) sequence was manually modified for generation of allele-specific primers targeting the aforesaid drug resistant mutations in the rpoB and katG gene of MTB genome. Specifically, for rpoB, D435V SNP position (GAC>GTC), H445D SNP position (CAC>GAC), S450L SNP position (TCG>TTG) and for katG, the S315T SNP position (AGC>ACC) was aligned to complement with the sequence of the FIP on its 3′ end. Nucleotide mismatches up to two bases were introduced at penultimate position on the FIP primer adjacent to SNP site of mutational allele. If the mutated nucleotide of the codon is designated "0," mismatches were introduced at position -1, or -2, of complementary FIP 3' end position in relation to the mutant-type (MT) sequence. The 3' terminus of each FIP was specific to the MT alleles responsible for conferring drug resistance. If the targeted location was an MT allele, the specific fragment was amplified otherwise, amplification would be prevented. The additional single-base mismatch at the -1 or -2 position from the 3' terminus of each forward allele-specific primer (FIP) was added to improve the discriminatory power between wild type (WT) and MT allele. Primer sets were screened via both the colorimetric and real-time fluorometric LAMP method, to ascertain their ability to discriminate between MT allele from the WT allele.

All primers were procured from Integrated DNA Technologies in lyophilized form and were dissolved in nuclease free ultrapure water (Invitrogen, Thermo Fisher Scientific) to a concentration of 100 µM and stored at -20 °C. 10X primer mix (1.6 µM

FIP & BIP, 0.4 µM LF & BF and 0.2 µM F3 & B3) was prepared for use in the LAMP assay.

**Designing and construction of plasmid positive control**

Plasmid templates were employed as positive control for the optimization of colorimetric and real-time LAMP assay. Genomic DNA samples of WT and MT (rpoB H445D and katG S315T) MTB were obtained from ICMR- NIRT, Chennai. All target genes were amplified by PCR using respective forward and backword primers yielding different amplicons which include IS6110 amplicons, IS1081 amplicons, full rpoB gene with H445D mutation (MT) & without any mutation (WT) and full katG gene with S315T mutation (MT) & without any mutation (WT). PCR products were purified by PCR Clean-up Kit (Promega, USA) and subsequently ligated with the pUC19 vector. The resulting recombinant plasmids were transformed into DH5-alpha cells followed by colony selection. Positive transformants were additionally cultured overnight at 37ºC in LB medium containing 100 mg/mL ampicillin. Then, plasmid DNA were isolated and purified using plasmid purification kit (Promega, USA). Plasmid constructs were confirmed by DNA Sanger sequencing. For constructing rpoB D435V and S450L mutant, site directed mutagenesis (SDM) was performed where specific mutation was introduced in WT plasmid. Primers were designed by QuikChange Primer Design software (Agilent Technologies). Mutation was incorporated by PCR amplification of the WT plasmids using PfuTurbo DNA polymerase enzyme (Agilent Technologies). Amplicons were subjected to the DpnI digestion and subsequently transformed into DH5-alpha cells. Colony selection, plasmid isolation and purification were performed as usual. The incorporated mutation was confirmed by the Sanger sequencing. The purified plasmids were diluted to prepare different working positive controls (Concentration from $1 \times 10^9$ to $1 \times 10$ copies/µL).

**Training Details of Staffs for working at PHC**

Few staffs for working at public health centres (PHC) were minimally trained using a structured, task-oriented protocol optimized for field-deployable molecular testing. A two months training comprised a brief theoretical orientation covering basic principles of isothermal amplification, biosafety, contamination control, and result interpretation, followed by intensive hands-on instruction. Practical training emphasized critical steps specific to LAMP workflows, including reagent reconstitution, template addition, maintenance of isothermal conditions, prevention of amplicon carryover, and interpretation of colorimetric or smartphone app-assisted readouts. Their competency was assessed using a composite evaluation framework encompassing theoretical understanding, direct observation of procedural adherence via structured checklists, concordance of test results with reference outcomes generated by trained personnel, and accuracy of result documentation. Quality assurance was maintained through periodic supervisory by project supervisors.

**Cohen's kappa (κ):**

$$k = \frac{P_o - P_e}{(1 + P_e)}$$

where,

$$P_o = \frac{TP + TN}{(TP + TN + FP + FN)}$$

$$P_e = \frac{(TP + FP)(TP + FN) + (FN + TN)(FP + TN)}{(TP + TN + FP + FN)^2}$$

$P_o$ = Observed agreement, $P_e$ = Expected agreement by chance, TP = True positive, TN = True negative, FP = False positive, FN = False negative

**Proportion of uninterpretable result:**

The proportion of uninterpretable result was calculated by the following formula as

$$Uninterpretable\ rate\ (\%) = \frac{Number\ of\ uninterpretable\ tests}{Total\ number\ of\ test\ performed} \times 100$$

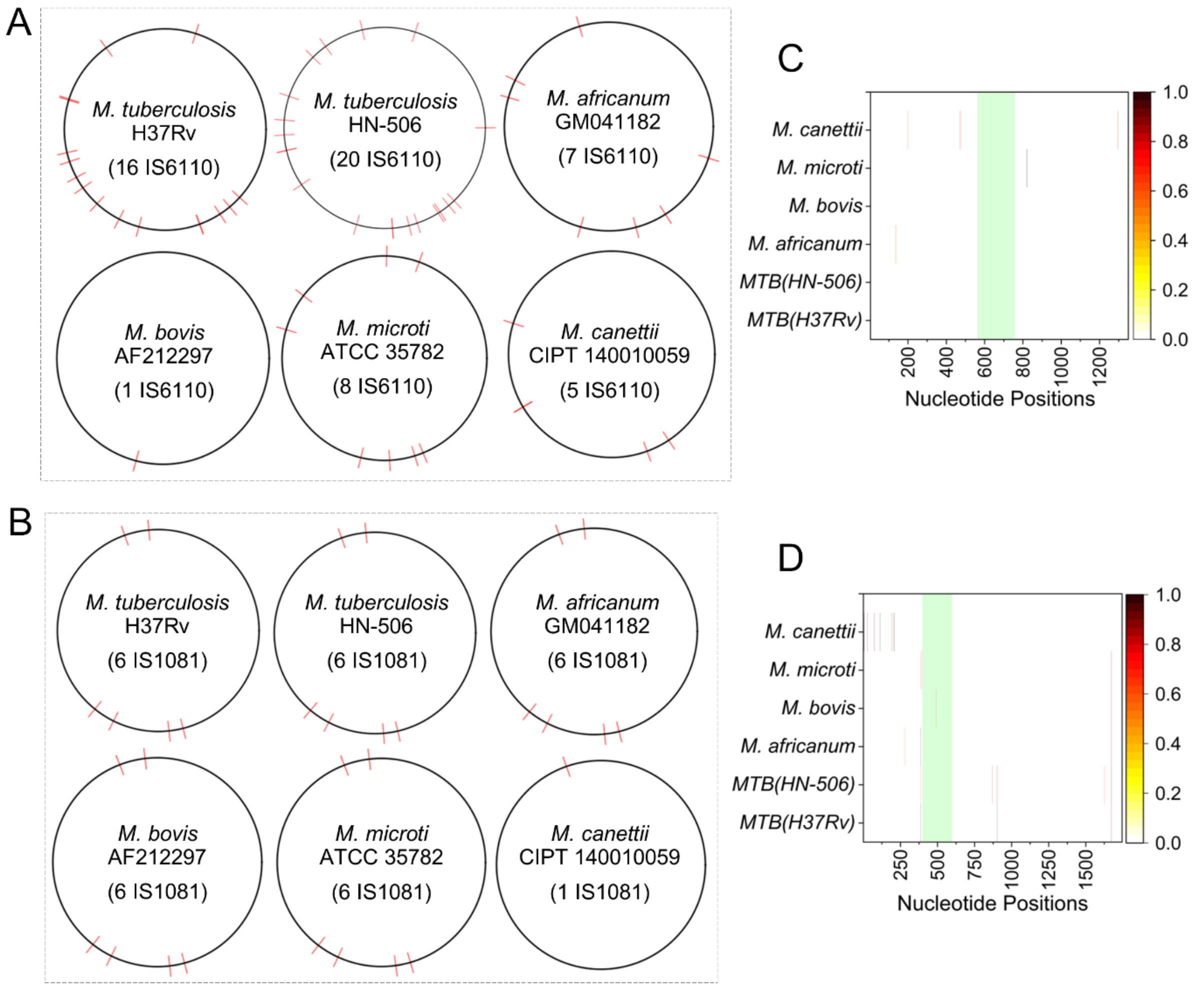


**Fig.S1.** CIRCOS map showing the position and number of (A) IS6110 and (B) IS1081 sequences in fully assembled genomes in indicated by red line. Heatmap graph showing the position of mutations in (C) IS6110 and (D) IS1081 sequences. The light green bar inside the graph indicate the target amplified positions.

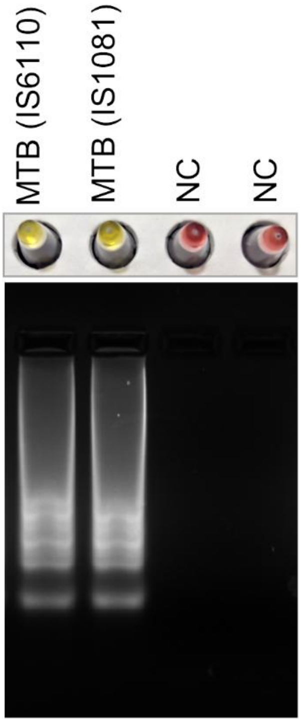


**Fig S2.** Positive sample showed band as typical ladder like pattern after successful LAMP reaction along with color changes to yellow. Negative samples showed no obvious color changes as well as typical band pattern.

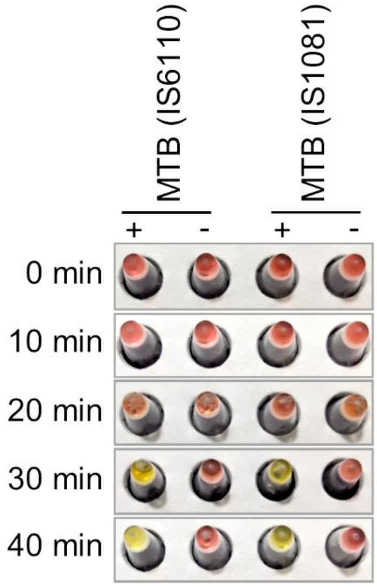


**Fig. S3.** Optimization of reaction time was performed at 65°C temperature on the basis of two set gene specific primer (IS6110 and IS1081). Typical yellow color changes in all sets were observed within 40 min.

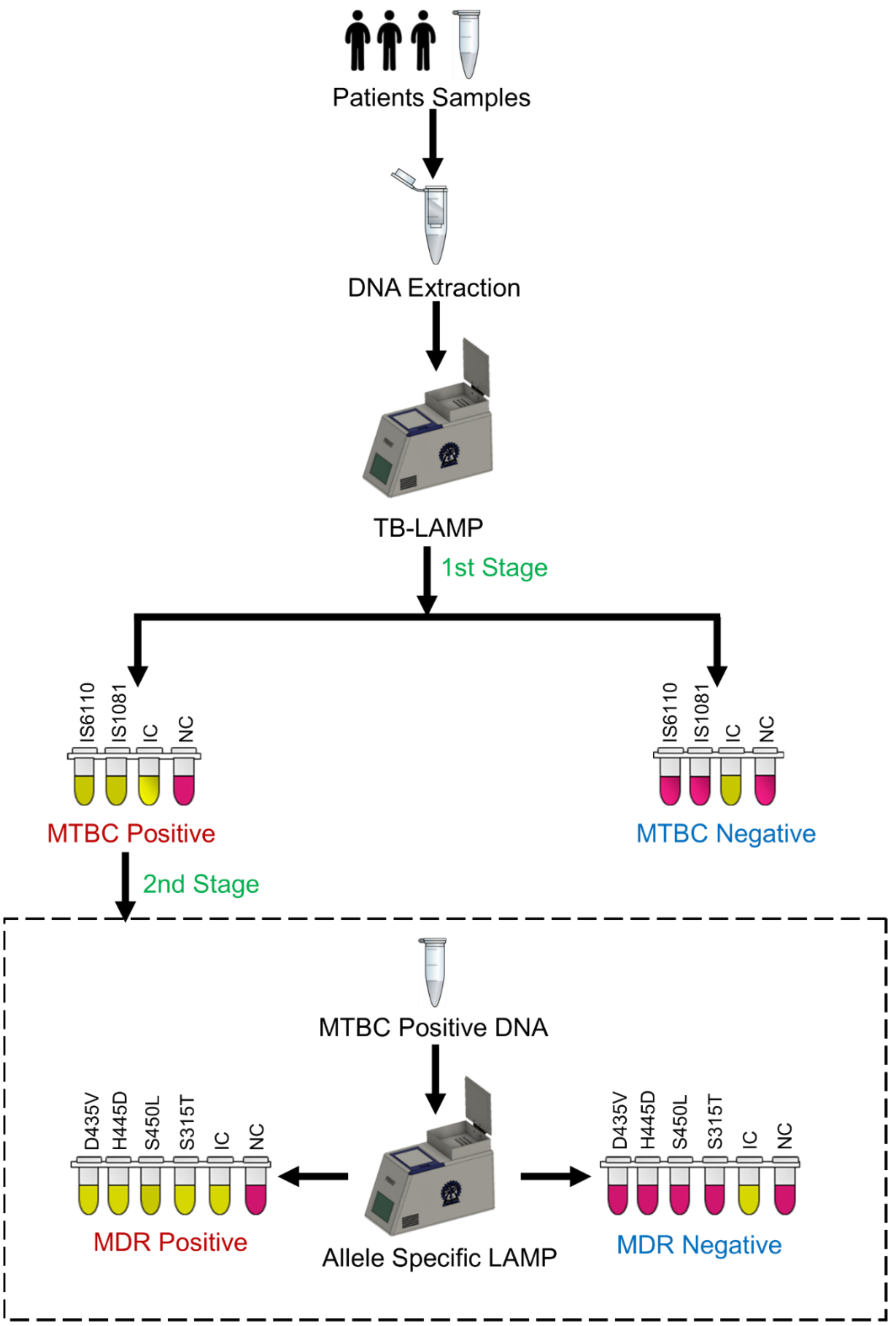


**Fig.S4:** Operational tiered testing workflow for TB with drug resistance. MTBC detection is performed as the primary assay and resistance assays will be performed only for the samples confirmed as MTBC-positive, not for the MTBC negative. Yellow color: Positive, Pink color: Negative IC: Internal control, NC: Negative control.

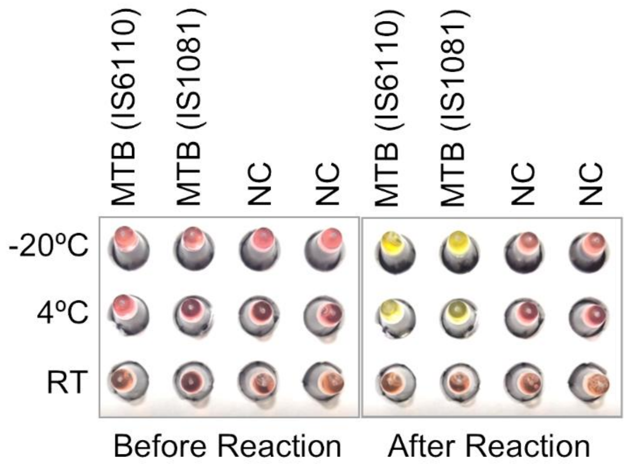


**Fig.S5.** Reagent stability characteristics for estimating the optimal storage temperature conditions. After reaction, yellow color for successful amplification, whereas pink color for unsuccessful amplification. PC stands for positive control, NC stands for negative control.

**Table S1.** Possible genetic mutations in IS6110 insertion sequence

| IS6110 | | | | | | | | | |
|---|---|---|---|---|---|---|---|---|---|
| Organism | Accession number | **% Identity** | **Region in genome** | | | **Region in query sequence** | | | **Mutations (Position in IS6110)** |
| | | | **Start** | **End** | **Length** | **Start** | **End** | **Length** | |
| **M. tuberculosis H37Rv** | AL123456.3 | 100 | 889034 | 890362 | 1329 | 1 | 1329 | 1329 | |
| | AL123456.3 | 100 | 1543293 | 1541965 | 1329 | 1 | 1329 | 1329 | |
| | AL123456.3 | 100 | 1989044 | 1987716 | 1329 | 1 | 1329 | 1329 | |
| | AL123456.3 | 100 | 1996114 | 1997442 | 1329 | 1 | 1329 | 1329 | |
| | AL123456.3 | 100 | 2365427 | 2366755 | 1329 | 1 | 1329 | 1329 | |
| | AL123456.3 | 100 | 2431458 | 2430130 | 1329 | 1 | 1329 | 1329 | |
| | AL123456.3 | 100 | 2550027 | 2551355 | 1329 | 1 | 1329 | 1329 | |
| | AL123456.3 | 100 | 2635590 | 2636918 | 1329 | 1 | 1329 | 1329 | |
| | AL123456.3 | 100 | 2785956 | 2784628 | 1329 | 1 | 1329 | 1329 | |
| | AL123456.3 | 100 | 2972122 | 2973450 | 1329 | 1 | 1329 | 1329 | |
| | AL123456.3 | 100 | 3121865 | 3120537 | 1329 | 1 | 1329 | 1329 | |
| | AL123456.3 | 100 | 3551243 | 3552571 | 1329 | 1 | 1329 | 1329 | |
| | AL123456.3 | 100 | 3552726 | 3554054 | 1329 | 1 | 1329 | 1329 | |
| | AL123456.3 | 100 | 3710395 | 3711723 | 1329 | 1 | 1329 | 1329 | |
| | AL123456.3 | 100 | 3796399 | 3795071 | 1329 | 1 | 1329 | 1329 | |
| | AL123456.3 | 100 | 3890792 | 3892120 | 1329 | 1 | 1329 | 1329 | |
| **M. tuberculosis HN-506** | AP018036.1 | 100 | 1608 | 2936 | 1329 | 1 | 1329 | 1329 | |
| | AP018036.1 | 100 | 887535 | 888863 | 1329 | 1 | 1329 | 1329 | |
| | AP018036.1 | 100 | 1262480 | 1261152 | 1329 | 1 | 1329 | 1329 | |
| | AP018036.1 | 100 | 1544967 | 1543639 | 1329 | 1 | 1329 | 1329 | |
| | AP018036.1 | 100 | 1659814 | 1658486 | 1329 | 1 | 1329 | 1329 | |
| | AP018036.1 | 100 | 1982186 | 1980858 | 1329 | 1 | 1329 | 1329 | |
| | AP018036.1 | 100 | 2151766 | 2150438 | 1329 | 1 | 1329 | 1329 | |
| | AP018036.1 | 100 | 2255539 | 2254211 | 1329 | 1 | 1329 | 1329 | |
| | AP018036.1 | 100 | 2362555 | 2363883 | 1329 | 1 | 1329 | 1329 | |
| | AP018036.1 | 100 | 2365613 | 2364285 | 1329 | 1 | 1329 | 1329 | |
| | AP018036.1 | 100 | 2624972 | 2626300 | 1329 | 1 | 1329 | 1329 | |
| | AP018036.1 | 100 | 3109266 | 3110594 | 1329 | 1 | 1329 | 1329 | |
| | AP018036.1 | 100 | 3363306 | 3361978 | 1329 | 1 | 1329 | 1329 | |
| | AP018036.1 | 100 | 3363810 | 3365138 | 1329 | 1 | 1329 | 1329 | |
| | AP018036.1 | 100 | 3481894 | 3480566 | 1329 | 1 | 1329 | 1329 | |
| | AP018036.1 | 100 | 3541077 | 3539749 | 1329 | 1 | 1329 | 1329 | |
| | AP018036.1 | 100 | 3704339 | 3703011 | 1329 | 1 | 1329 | 1329 | |
| | AP018036.1 | 100 | 3718657 | 3719985 | 1329 | 1 | 1329 | 1329 | |
| | AP018036.1 | 100 | 3794236 | 3795564 | 1329 | 1 | 1329 | 1329 | |
| | AP018036.1 | 100 | 3844797 | 3843469 | 1329 | 1 | 1329 | 1329 | |
| **M. africanum GM041182** | FR878060.1 | 100 | 1298913 | 1297585 | 1329 | 1 | 1329 | 1329 | |
| | FR878060.1 | 100 | 1873018 | 1874346 | 1329 | 1 | 1329 | 1329 | |
| | FR878060.1 | 100 | 1996349 | 1997677 | 1329 | 1 | 1329 | 1329 | |
| | FR878060.1 | 100 | 3474729 | 3473401 | 1329 | 1 | 1329 | 1329 | |
| | FR878060.1 | 100 | 3691507 | 3690179 | 1329 | 1 | 1329 | 1329 | |

| | | | | | | | | | |
|---|---|---|---|---|---|---|---|---|---|
| | FR878060.1 | 100 | 4174642 | 4175970 | 1329 | 1 | 1329 | 1329 | |
| | FR878060.1 | 99.925 | 3099319 | 3097991 | 1329 | 1 | 1329 | 1329 | 1(A137G) |
| **M. bovis AF212297** | LT708304.1 | 100 | 3082757 | 3081429 | 1329 | 1 | 1329 | 1329 | |
| **M.canettii CIPT 140010059** | NC_015848.1 | 99.925 | 3609840 | 3611168 | 1329 | 1 | 1329 | 1329 | 1(G1296A) |
| | NC_015848.1 | 99.85 | 2023455 | 2022127 | 1329 | 1 | 1329 | 1329 | 2(G200A, G473A) |
| | NC_015848.1 | 100 | 3779421 | 3778262 | 1160 | 1 | 1160 | 1160 | 0 |
| | NC_015848.1 | 100 | 2621524 | 2622305 | 782 | 548 | 1329 | 782 | 0 |
| | NC_015848.1 | 100 | 2619699 | 2620249 | 551 | 1 | 551 | 551 | 0 |
| **M. microti ATCC 35782** | LR882496.1 | 99.925 | 854647 | 853319 | 1329 | 1 | 1329 | 1329 | 1 (G821T) |
| | LR882496.1 | 99.925 | 1079401 | 1078073 | 1329 | 1 | 1329 | 1329 | 1 (G821T) |
| | LR882496.1 | 99.925 | 1726336 | 1725008 | 1329 | 1 | 1329 | 1329 | 1 (G821T) |
| | LR882496.1 | 99.925 | 1986424 | 1985096 | 1329 | 1 | 1329 | 1329 | 1 (G821T) |
| | LR882496.1 | 99.925 | 3111662 | 3110334 | 1329 | 1 | 1329 | 1329 | 1 (G821T) |
| | LR882496.1 | 99.925 | 3307118 | 3308446 | 1329 | 1 | 1329 | 1329 | 1 (G821T) |
| | LR882496.1 | 99.925 | 3483517 | 3482189 | 1329 | 1 | 1329 | 1329 | 1 (G821T) |
| | LR882496.1 | 99.925 | 3541815 | 3543143 | 1329 | 1 | 1329 | 1329 | 1 (G821T) |

**Table S2.** Possible genetic mutations in IS1081 insertion sequence

| IS1081 | | | | | | | | | |
|---|---|---|---|---|---|---|---|---|---|
| **Organism** | **Accession number** | **% Identity** | **Region in genome** | | | **Region in query sequence** | | | **Mutations (Position in IS1081)** |
| | | | **Start** | **End** | **Length** | **Start** | **End** | **Length** | |
| **M. tuberculosis H37Rv** | AL123456.3 | 100 | 1342966 | 1341287 | 1680 | 1 | 1680 | 1680 | 0 |
| | AL123456.3 | 99.93 | 2829928 | 2828491 | 1438 | 237 | 1674 | 1438 | 1 (T1672C) |
| | AL123456.3 | 99.861 | 1169298 | 1170732 | 1435 | 237 | 1671 | 1435 | 2 (A386G, A904G) |
| | AL123456.3 | 99.861 | 3382747 | 3381313 | 1435 | 237 | 1671 | 1435 | 2 (A386G, A904G) |
| | AL123456.3 | 99.861 | 3481326 | 3482760 | 1435 | 237 | 1671 | 1435 | 2 (A386G, A904G) |
| | AL123456.3 | 99.89 | 2982946 | 2983854 | 909 | 237 | 1145 | 909 | 1 (A904G) |
| **M. tuberculosis HN-506** | AP018036.1 | 99.94 | 1342500 | 1340821 | 1680 | 1 | 1680 | 1680 | 1 (A386G) |
| | AP018036.1 | 99.861 | 1167318 | 1168752 | 1435 | 237 | 1671 | 1435 | 2(A386G, A904G) |
| | AP018036.1 | 99.791 | 2820093 | 2818656 | 1438 | 237 | 1674 | 1438 | 3 (A386G, A904G, T1672C) |
| | AP018036.1 | 99.861 | 3368875 | 3367441 | 1435 | 237 | 1671 | 1435 | 2 (A386G, A904G) |
| | AP018036.1 | 99.791 | 3467968 | 3469402 | 1435 | 237 | 1671 | 1435 | 3(A386G, A904G, A1628G) |
| | AP018036.1 | 99.89 | 2971746 | 2972654 | 909 | 237 | 1145 | 909 | 1 (A386G) |
| **M. africanum GM041182** | FR878060.1 | 99.94 | 1341701 | 1340022 | 1680 | 1 | 1680 | 1680 | 1 (G280A) |
| | FR878060.1 | 100 | 3361687 | 3360253 | 1435 | 237 | 1671 | 1435 | 0 |
| | FR878060.1 | 100 | 3460321 | 3461755 | 1435 | 237 | 1671 | 1435 | 0 |
| | FR878060.1 | 99.93 | 1165695 | 1167129 | 1435 | 237 | 1671 | 1435 | 1 (A386G) |
| | FR878060.1 | 99.861 | 2816716 | 2815279 | 1438 | 237 | 1674 | 1438 | 2(A386G, T1672C) |
| | FR878060.1 | 99.89 | 2959410 | 2960318 | 909 | 237 | 1145 | 909 | 1(G280A) |
| **M. bovis AF212297** | LT708304.1 | 99.94 | 1344241 | 1342562 | 1680 | 1 | 1680 | 1680 | 1(490T) |
| | LT708304.1 | 100 | 1169775 | 1171209 | 1435 | 237 | 1671 | 1435 | 0 |
| | LT708304.1 | 99.93 | 2801136 | 2799699 | 1438 | 237 | 1674 | 1438 | 1 (T1672C) |
| | LT708304.1 | 100 | 3343719 | 3342285 | 1435 | 237 | 1671 | 1435 | 0 |
| | LT708304.1 | 100 | 3442285 | 3443719 | 1435 | 237 | 1671 | 1435 | 0 |
| | LT708304.1 | 100 | 2943837 | 2944745 | 909 | 237 | 1145 | 909 | 0 |
| **M. canettii CIPT 140010059** | NC_015848.1 | 99.643 | 1363022 | 1361343 | 1680 | 1 | 1680 | 1680 | 6 (T5C, G28T, C73G, A112G, A195G, G208T) |
| **M. microti ATCC 35782** | LR882496.1 | 100 | 1346001 | 1344322 | 1680 | 1 | 1680 | 1680 | 0 |
| | LR882496.1 | 100 | 3370599 | 3369165 | 1435 | 237 | 1671 | 1435 | 0 |
| | LR882496.1 | 100 | 3469233 | 3470667 | 1435 | 237 | 1671 | 1435 | 0 |
| | LR882496.1 | 99.93 | 1173592 | 1175026 | 1435 | 237 | 1671 | 1435 | 1(G394T) |
| | LR882496.1 | 99.861 | 2821978 | 2820541 | 1438 | 237 | 1674 | 1438 | 2(A386G, T1672C) |
| | LR882496.1 | 100 | 2973678 | 2974586 | 909 | 237 | 1145 | 909 | 0 |

**Table S3.** Result of assay precision for detecting TB targeting IS6110

| Bacterial load (copies/µL) | Threshold Time (Tt) | | | | SD | CV (%) |
|---|---|---|---|---|---|---|
| | Run 1 | Run 2 | Run 3 | Mean | | |
| $1.0\times10^{1}$ | 29.15 | 29.42 | 29.63 | 29.4 | 0.196 | 0.668 |
| $1.0\times10^{2}$ | 24.07 | 23.97 | 24.31 | 24.12 | 0.143 | 0.592 |
| $1.0\times10^{3}$ | 20.03 | 19.89 | 20.25 | 20.06 | 0.148 | 0.739 |
| $1.0\times10^{4}$ | 16.65 | 16.23 | 16.89 | 16.59 | 0.273 | 1.64 |
| $1.0\times10^{5}$ | 12.98 | 12.5 | 13.2 | 12.89 | 0.292 | 2.267 |
| $1.0\times10^{6}$ | 9.9 | 10.21 | 9.78 | 9.96 | 0.181 | 1.818 |

**Table S4.** Result of assay precision for detecting TB targeting IS1081

| Bacterial load (copies/µL) | Threshold Time (Tt) | | | | SD | CV (%) |
|---|---|---|---|---|---|---|
| | Run 1 | Run 2 | Run 3 | Mean | | |
| $1.0\times10^{1}$ | 30.99 | 31.4 | 30.71 | 31.03 | 0.283 | 0.913 |
| $1.0\times10^{2}$ | 24.2 | 24.6 | 23.93 | 24.24 | 0.275 | 1.135 |
| $1.0\times10^{3}$ | 19.98 | 19.51 | 20.1 | 19.86 | 0.255 | 1.282 |
| $1.0\times10^{4}$ | 13.61 | 13.1 | 13.83 | 13.51 | 0.305 | 2.263 |
| $1.0\times10^{5}$ | 10.7 | 10.2 | 10.95 | 10.62 | 0.312 | 2.937 |
| $1.0\times10^{6}$ | 7.7 | 7.98 | 8.1 | 7.93 | 0.167 | 2.114 |

**Table S5:** Details of patient's samples used in field evaluation study and their overall outcome (without 'uninterpreted result')

| Sample ID | Gender | Sample Type | GeneXpert | TB-LAMP | Remarks |
|---|---|---|---|---|---|
| T1 | Male | Sputum | Positive | Positive | Concordant |
| T2 | Male | Sputum | Positive | Positive | Concordant |
| T3 | Female | Sputum | Positive | Positive | Concordant |
| T4 | Male | Sputum | Positive | Positive | Concordant |
| T5 | Male | Sputum | Positive | Positive | Concordant |
| T6 | Male | Sputum | Negative | Negative | Concordant |
| T7 | Male | Sputum | Negative | Negative | Concordant |
| T8 | Male | Sputum | Positive | Positive | Concordant |
| T9 | Male | Sputum | Negative | Negative | Concordant |
| T10 | Male | Sputum | Negative | Negative | Concordant |
| T11 | Female | Sputum | Positive | Positive | Concordant |
| T12 | Female | Sputum | Positive | Positive | Concordant |
| T13 | Female | Sputum | Positive | Positive | Concordant |
| T14 | Female | Sputum | Positive | Negative | Non-concordant |
| T15 | Female | Sputum | Positive | Positive | Concordant |
| T16 | Male | Sputum | Positive | Positive | Concordant |
| T17 | Male | Sputum | Positive | Positive | Concordant |
| T18 | Male | Sputum | Negative | Negative | Concordant |
| T19 | Male | Sputum | Positive | Positive | Concordant |
| T20 | Male | Sputum | Positive | Negative | Non-concordant |
| T21 | Male | Sputum | Negative | Negative | Concordant |
| T22 | Male | Sputum | Positive | Positive | Concordant |
| T23 | Female | Sputum | Positive | Positive | Concordant |
| T24 | Female | Sputum | Positive | Positive | Concordant |
| T25 | Female | Sputum | Positive | Positive | Concordant |
| T26 | Male | Sputum | Positive | Negative | Non-concordant |
| T27 | Male | Sputum | Positive | Positive | Concordant |
| T28 | Female | Sputum | Positive | Positive | Concordant |
| T29 | Female | Sputum | Positive | Positive | Concordant |
| T30 | Male | Sputum | Positive | Positive | Concordant |
| T31 | Male | Sputum | Negative | Negative | Concordant |
| T32 | Male | Sputum | Negative | Negative | Concordant |
| T33 | Female | Sputum | Positive | Positive | Concordant |
| T34 | Female | Sputum | Positive | Positive | Concordant |
| T35 | Male | Sputum | Positive | Positive | Concordant |
| T36 | Male | Sputum | Positive | Positive | Concordant |
| T37 | Female | Sputum | Positive | Positive | Concordant |
| T38 | Female | Sputum | Negative | Negative | Concordant |
| T39 | Female | Sputum | Negative | Negative | Concordant |
| T40 | Male | Sputum | Negative | Negative | Concordant |

| | | | | | |
|---|---|---|---|---|---|
| T41 | Male | Sputum | Positive | Positive | Concordant |
| T42 | Male | Sputum | Positive | Positive | Concordant |
| T43 | Male | Sputum | Positive | Positive | Concordant |
| T44 | Male | Sputum | Positive | Positive | Concordant |
| T45 | Male | Sputum | Positive | Positive | Concordant |
| T46 | Male | Sputum | Negative | Negative | Concordant |
| T47 | Female | Sputum | Positive | Positive | Concordant |
| T48 | Male | Sputum | Positive | Positive | Concordant |
| T49 | Male | Sputum | Positive | Positive | Concordant |
| T50 | Female | Sputum | Positive | Positive | Concordant |
| T51 | Female | Sputum | Positive | Positive | Concordant |
| T52 | Male | Sputum | Positive | Positive | Concordant |
| T53 | Female | Sputum | Positive | Positive | Concordant |
| T54 | Male | Sputum | Positive | Positive | Concordant |
| T55 | Male | Sputum | Positive | Positive | Concordant |
| T56 | Female | Sputum | Positive | Positive | Concordant |
| T57 | Male | Sputum | Positive | Positive | Concordant |
| T58 | Male | Sputum | Negative | Negative | Concordant |
| T59 | Male | Sputum | Positive | Positive | Concordant |
| T60 | Female | Sputum | Negative | Negative | Concordant |
| T61 | Female | Sputum | Negative | Negative | Concordant |
| T62 | Male | Sputum | Positive | Positive | Concordant |
| T63 | Male | Sputum | Positive | Positive | Concordant |
| T64 | Male | Sputum | Positive | Positive | Concordant |
| T65 | Male | Sputum | Positive | Negative | Non-concordant |
| T66 | Male | Sputum | Positive | Positive | Concordant |
| T67 | Male | Sputum | Positive | Positive | Concordant |
| T68 | Female | Sputum | Negative | Negative | Concordant |
| T69 | Female | Sputum | Negative | Negative | Concordant |
| T70 | Male | Sputum | Negative | Negative | Concordant |
| T71 | Female | Sputum | Negative | Negative | Concordant |
| T72 | Female | Sputum | Positive | Positive | Concordant |
| T73 | Male | Sputum | Positive | Positive | Concordant |
| T74 | Male | Sputum | Positive | Positive | Concordant |
| T75 | Female | Sputum | Positive | Positive | Concordant |
| T76 | Male | Sputum | Positive | Positive | Concordant |
| T77 | Female | Sputum | Positive | Positive | Concordant |
| T78 | Female | Sputum | Positive | Positive | Concordant |
| T79 | Male | Sputum | Positive | Positive | Concordant |
| T80 | Male | Sputum | Positive | Positive | Concordant |
| T81 | Male | Sputum | Positive | Positive | Concordant |
| T82 | Male | Sputum | Negative | Negative | Concordant |
| T83 | Male | Sputum | Negative | Negative | Concordant |
| T84 | Female | Sputum | Positive | Positive | Concordant |
| T85 | Male | Sputum | Positive | Positive | Concordant |
| T86 | Male | Sputum | Positive | Positive | Concordant |

| | | | | | |
|---|---|---|---|---|---|
| T87 | Male | Sputum | Positive | Positive | Concordant |
| T88 | Male | Sputum | Positive | Positive | Concordant |
| T89 | Male | Sputum | Positive | Positive | Concordant |
| T90 | Male | Sputum | Negative | Negative | Concordant |
| T91 | Male | Sputum | Negative | Negative | Concordant |
| T92 | Female | Sputum | Positive | Positive | Concordant |
| T93 | Male | Sputum | Positive | Positive | Concordant |
| T94 | Male | Sputum | Positive | Positive | Concordant |
| T95 | Female | Sputum | Positive | Positive | Concordant |
| T96 | Male | Sputum | Positive | Positive | Concordant |
| T97 | Female | Sputum | Positive | Positive | Concordant |
| T98 | Male | Sputum | Positive | Positive | Concordant |
| T99 | Female | Sputum | Negative | Negative | Concordant |
| T100 | Male | Sputum | Negative | Negative | Concordant |
| T101 | Male | Sputum | Positive | Positive | Concordant |
| T102 | Male | Sputum | Positive | Positive | Concordant |
| T103 | Male | Sputum | Positive | Positive | Concordant |
| T104 | Female | Sputum | Positive | Positive | Concordant |
| T105 | Female | Sputum | Positive | Positive | Concordant |
| T106 | Female | Sputum | Positive | Positive | Concordant |
| T107 | Male | Sputum | Negative | Negative | Concordant |
| T108 | Male | Sputum | Negative | Negative | Concordant |
| T109 | Male | Sputum | Positive | Positive | Concordant |
| T110 | Female | Sputum | Positive | Positive | Concordant |
| T111 | Male | Sputum | Positive | Positive | Concordant |
| T112 | Female | Sputum | Positive | Positive | Concordant |
| T113 | Male | Sputum | Positive | Positive | Concordant |
| T114 | Female | Sputum | Negative | Negative | Concordant |
| T115 | Male | Sputum | Positive | Positive | Concordant |
| T116 | Female | Sputum | Positive | Positive | Concordant |
| T117 | Female | Sputum | Positive | Positive | Concordant |
| T118 | Male | Sputum | Positive | Positive | Concordant |
| T119 | Female | Sputum | Positive | Positive | Concordant |

**Table S6:** Technology wise comparison of TB-LAMP assay, GeneXpert and Truenat

| Key Features | TB-LAMP | GeneXpert | Truenat |
|---|---|---|---|
| USP | Fast & highly sensitive molecular test for detection of MTB infection directly from sputum samples/extracted DNA implementable at POC. | Accurate molecular test for detection of MTB infection from sputum samples. | Fast & accurate molecular test for detection of MTB infection from sputum samples. |
| Principle for detection | Isothermal Nucleic acid amplification (LAMP based) | PCR based nucleic amplification. | PCR based nucleic amplification. |
| Prior DNA extraction | Required | Through chip based | Required through automated extraction machine |
| Sensitivity | 50-100 CFU/ml | 100 CFU/ml | 100 CFU/ml |
| Cost per test (USD) | ~3.0 | ~40.0 | ~10.0 |
| Assay run time | 40 min | 2 hours | 1 hour |
| Ease of use | Minimally trained technician needed | Highly trained technician needed | High to moderate trained technician needed |
| Laboratory facility required | Extreme point of care settings with limited resources | Need uninterrupted electric supply and air-conditioning for proper functioning | Point of care settings with limited resources |
| Mode of detection | Colorimetric/Fluorescence curve | Value/Data | Value/Data |

### REFERENCES


1. Zaw MT, Emran NA, Lin Z. 2018. Mutations inside rifampicin-resistance determining region of rpoB gene associated with rifampicin-resistance in Mycobacterium tuberculosis. Journal of Infection and Public Health 11:605–610.
2. Indian Catalogue of *Mycobacterium tuberculosis* Mutations and their Association with Drug Resistance Version 2.0. 2024. ICMR-National Institute for Research in Tuberculosis.
3. Catalogue of mutations in Mycobacterium tuberculosis complex and their association with drug resistance, second edition. Geneva: World Health Organization; 2023. Licence: CC BY-NC-SA 3.0 IGO.
